# Serotype-specific pneumococcal invasiveness: a global meta-analysis of paired estimates of disease incidence and carriage prevalence

**DOI:** 10.1101/2025.03.03.25323223

**Authors:** Katherine E. Gallagher, Fredrick Odiwour, Christian Bottomley, John Ojal, Aisha Adamu, Esther Muthumbi, Eunice W. Kagucia, Laura L Hammitt, Sergio Massora, Betuel Sigaúque, Alberto Chaúque, Leocadia Vilanculos, Jennifer R. Verani, Maria da Gloria Carvalho, Anne von Gottberg, Jackie Kleynhans, Shabir A. Madhi, Courtney P. Olwagen, Grant Mackenzie, Rasheed Salaudeen, Ryan Gierke, Miwako Kobayashi, Stephen Pelton, Inci Yildirim, Stepy Thomas, Amy Tunali, Monica Farley, Todd D. Swarthout, Akuzike Kalizang’oma, Robert S. Heyderman, Neil French, Yoon Choi, Nick Andrews, Shamez Ladhani, Elizabeth Miller, J. Anthony G. Scott

## Abstract

**Background:** Serotype-specific estimates of pneumococcal invasiveness used in pneumococcal carriage transmission models to predict changes in disease incidence post-vaccination are largely derived from high-income settings. We conducted a systematic review of carriage prevalence and invasive pneumococcal disease (IPD) incidence to calculate case-carrier ratios (CCRs) in different income settings.

**Methods:** A systematic search of Medline, Embase, and Global Health databases in March 2022 identified publications on pneumococcal carriage prevalence or IPD incidence; we requested individual-level data from authors of relevant texts. Serotype-specific CCRs, calculated as IPD incidence divided by carriage prevalence, were pooled across settings using random effects meta-analyses, stratified by pre-/post-pneumococcal conjugate vaccine (PCV) introduction, country income group, age-group, sex and HIV status.

**Findings:** We identified 80 publications from 18 countries (13 upper-middle- or high-income countries (UM/HIC), 5 low/lower-middle income (L/LMIC)) reporting carriage prevalence or IPD incidence in overlapping geographical areas, time periods, and age-groups. We calculated CCRs for >70 serotypes, stratified by age group, income settings, and pre- and post-vaccine introduction. In children under five, pre-PCV CCRs for serotypes not included in the 20-valent PCV were higher in L/LMICs than UM/HICs, 152 (95% Confidence interval 103-226) versus 102 (50-209). Post-PCV CCRs for non-vaccine serotypes dropped in UM/HICs but not in L/LMICs, 19 (16-22) versus 154 (119-200) respectively. Pre-/post PCV changes varied by serotype and age-group. CCRs were lowest in 5–14-year-olds and were higher in HIV positive than HIV negative individuals. There were no differences in CCRs by sex.

**Interpretation:** Pneumococcal invasiveness varies by serotype, age-group, country income-group, HIV status and over time; however, substantial variation remained unexplained. Our CCRs represent the most representative estimates of invasiveness currently available for use in statistical or mathematical prediction models of disease incidence, where only carriage prevalence data are available.

**Funding:** The Wellcome Trust, Great Britain (098532)

**Panel: Research in context:** *Evidence before this study:* There are three estimates of the absolute risk of invasive pneumococcal disease, given carriage, derived from data from high-income settings (two studies in the UK, and one in the USA). A fourth set of estimates have been derived from data collated by a recent review of studies that reported both carriage and IPD data in the same publication. This review and re-analysis combined data from 12 countries to report case-carrier ratios in children under-5, pre- and post-vaccine introduction. The review did not include data from IPD surveillance sites in low- and middle-income countries, nor carriage prevalence data in adults.

*Added value of this study:* We conducted an extensive systematic review to identify high quality IPD incidence estimates and a comprehensive database of carriage prevalence estimates that arise from the same country, age-group and time period as these IPD incidence estimates. We employed stringent matching criteria to only include the results of carriage surveys that were conducted in a random sample of the general population, and IPD surveillance activities that were conducted in a systematic way across a defined population. This enabled us to estimate serotype-specific pneumococcal case-carrier ratios, stratified by age group, country income group, and time period pre- or post-vaccine introduction.

*Implications of all the available evidence:* Invasive pneumococcal disease surveillance is resource intensive to establish and sustain and is therefore infeasible for most countries worldwide. Pneumococcal vaccine policy is often made on the basis of carriage data alone, or mathematical models which predict changes in disease incidence by combining changes in carriage prevalence with pre-specified case-carrier ratios. We have used all available data globally to estimate serotype-specific case-carrier ratios, which previously have been derived from data from high income settings. Both statistical and mathematical models predicting changes in disease incidence in low-income settings, can now utilise case-carrier ratios from more relevant population groups. This will be of increasing importance as policy makers attempt to make evidence-based decisions on whether to change pneumococcal vaccine product, schedule, or simply increase coverage of the existing programme.

## Introduction

*Streptococcus pneumoniae* (pneumococcus) causes respiratory tract infections, sepsis, and meningitis. In 2021, pneumococci are estimated to have caused 98 million episodes of lower respiratory tract infection (uncertainty interval, UI, 92–104) and 5051000 deaths (UI 4541000– 5551000) worldwide(1). Together with bacteraemia and meningitis, this equates to 493 disability-adjusted life years (DALYs) lost per 100,000 population across all ages (UI 408-602)(2).

Carriage of pneumococci is a necessary precursor to disease(3). Pneumococcal conjugate vaccines (PCVs) have been highly effective at preventing carriage and disease caused by the pneumococcal serotypes included in the vaccine (‘vaccine types’, VTs), but the vaccines are expensive(4). New PCVs which expand protection of the current 10 and 13-valent products, to 15 or 20 serotypes, have been recently licensed for paediatric use by the European Medicines Agency and other higher valency products are in development. Evidence of PCV impact on disease is essential to justify the PCV programme’s ongoing expense and predict the impact of future changes in vaccine schedule or product(5).

PCV impact studies would ideally use invasive pneumococcal disease (IPD) surveillance as the primary outcome(6); however, IPD surveillance is resource intensive to establish and maintain over the long periods of time needed to measure changes in incidence, and to then attribute these changes to a particular vaccine policy(7). Carriage surveys are easier and less costly to conduct, but the outputs then need to be translated into a clinically relevant outcome, like IPD. Accurate measures of serotype-specific pneumococcal invasiveness could be used in statistical models to convert carriage prevalence estimates from carriage surveys into an estimate of disease burden in areas where IPD surveillance is too costly to implement(8, 9). Mathematical models of the impact of new PCVs already use estimates of invasiveness (case-carrier ratios [CCRs]) to predict the impact of the replacement in carriage of vaccine types with non-vaccine types on disease burden(10).

Prior estimates of invasiveness have demonstrated that invasiveness varies significantly by serotype(11, 12). In high-income settings, the absolute risk of invasion, given carriage, has been estimated using data on carriage prevalence and IPD incidence (13–15). A recent review(16) re-analysed data from studies that reported both carriage prevalence and IPD incidence in order to calculate CCR, but did not include data from IPD surveillance sites in low and middle income countries, nor carriage prevalence data from adults(17–19). We aimed to estimate serotype-specific pneumococcal CCRs by collating all available data on carriage prevalence and IPD incidence globally, and to determine if invasiveness varies by age, sex, HIV status, country income group, and pre- or post-PCV introduction, to inform the use of CCRs in estimating IPD burden.

## Methods

### Search Strategy and selection criteria

We conducted systematic searches of Medline, Embase (Ovid SP), and Global Heath databases, for publications and conference abstracts using search terms for pneumococcus, and carriage or invasive disease (**Supplementary Figure 1**). Searches were performed on 14^th^ March 2022 with no geographical, age, or year restrictions. Titles and abstracts were screened by two authors. Titles and abstracts that reported any pneumococcal carriage or disease incidence data, or both, were included if an abstract was available in English. Full texts were retrieved, screened by two authors, and included in a long-list if they reported the serotype-specific results of either representative surveys of carriage prevalence among healthy people, or IPD surveillance activities in a defined population (**Figure 1**). Discrepancies in selection were resolved by a third author.

**Figure 1:**
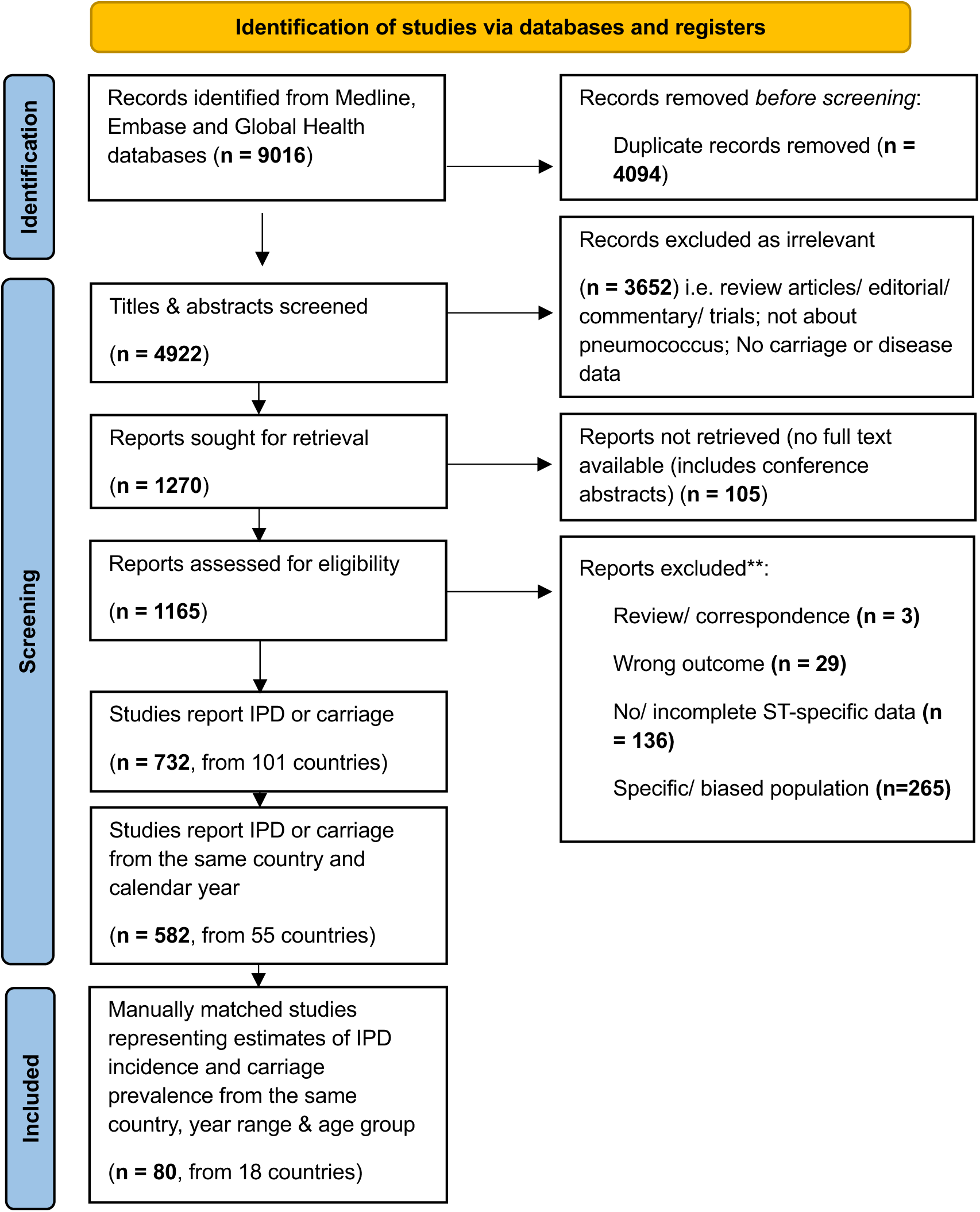
PRISMA flow diagram for the systematic review of the literature.

To further refine the selection, one author extracted basic study characteristics from all full texts i.e., the outcome reported, calendar year(s), country(ies) and age-groups included in data collection. A process of matching reports of carriage prevalence and IPD incidence was then undertaken. Publications of carriage prevalence were included if they overlapped by at least one calendar year with data available on IPD incidence from the same country. In addition, both carriage and IPD data had to have been collected in the same PCV period (either pre-PCV or post-PCV). Publications on IPD and carriage in the same country and calendar period had to have sampled within the same age group (i.e., aged under-5-years (data reported in children less than 6 or 7-years were also included), 5-14 year olds (data reported in 5-17 or 5-18 year olds were also included in this group), and 15 years and above (data reported in 18 year olds and above were also included in this group)). Additionally, if both carriage and IPD data were from children aged under-5-years, both estimates of carriage and IPD had to either include or exclude infants (aged <12 months). If estimates of carriage and IPD were both in 15-year-olds and above, both estimates had to either include or exclude those aged >65-years. This is because data from infants and >65s dominate IPD incidence estimates.

### Data extraction

Investigators were approached to provide individual level data on IPD cases, population denominators and carriage survey participants, including, where available, age, sex, PCV, and HIV status. Data were shared by investigators from Kenya, Malawi, Mozambique, South Africa, The Gambia, United States of America and England and Wales. If individual level data could not be obtained (an additional 11 countries), data were extracted from publications. The country’s World Bank income group was based on the classifications available in 2022. PCV introduction status at the time of sampling was defined using data from VIEW-hub(20).

### Statistical analysis

For each serotype-specific ‘match’ of IPD incidence data to carriage prevalence data, a case-carrier ratio (CCR) and an estimate of its variance was calculated using formula:

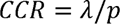

where *λ* is the incidence of IPD, calculated as the number of cases of IPD per 100,000 person-years, and *p* is the carriage prevalence. A 95% confidence interval for each CCR was obtained via the delta method(21) (**Supplementary Figure 2**).

Where there was an estimate of carriage prevalence for a specific serotype, age and year group, but there were zero reported cases of disease, or, where there were reported cases of disease but zero detected carriers, a case-carrier ratio could not be computed. To avoid underestimating the invasiveness of highly invasive but rarely carried serotypes (e.g., serotypes 1 and 5) and overestimating the invasiveness of frequently carried but less invasive serotypes, we replaced zero incidence and prevalence estimates with imputed small, non-zero values. Imputation used an empirical Bayes approach. We fitted serotype-specific beta binomial models to the observed counts, with age-group, country income group and PCV introduction status as covariates(22). The beta distribution estimated from this model was used as a prior and combined with the observed zero count to derive a posterior distribution. The mean of this posterior, *µ_post_*, was used to generate a small non-zero imputed value:

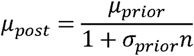

where *µ_prior_* and *a_prior_* are, respectively, the mean and overdispersion parameter of the beta prior (estimated by fitting the beta-binomial model) and *n* is the sample size (carriage prevalence) or person years (IPD incidence) of the study. Note that the imputed value, *µ_post_*, is close to the µ_prior_ when n is close to zero and close to zero when n is large. The resultant imputed values were only used if observed counts were zero.

To combine serotype-specific CCRs across studies and groups, summary estimates were produced using serotype-specific random effects meta-analyses, allowing for estimates to vary between studies(23). Summary estimates were stratified by age, country income group, and calendar year or period (pre- or post-PCV) and were made available for download via a Shiny app (https://pneumovaxpol.shinyapps.io/Pneumo_CCR_MetaAnalysis/).

In internal calibration, reported serotype-specific carriage prevalences post-PCV introduction were multiplied by the serotype-specific CCR estimate obtained by pooling data from all the countries in the same income group to calculate the predicted IPD incidence in under-5s. This was compared with observed IPD incidence. Exploratory analyses of the variation in invasiveness across countries used country level data extracted from the UNAID and WHO Global Health Observatory databases, including HIV prevalence in adults aged 15-49 years, the prevalence of underweight children, the proportion of children with pneumonia who are taken to a health facility, the prevalence of handwashing facilities and cooking with biomass at home, and third dose PCV coverage among infants. A mixed linear effects model was built to assess the association of these country-level factors with CCRs, controlling for clustering by serotype.

The individual level data obtained from authors were split into datasets corresponding to sex (male or female) and HIV status (infected or uninfected). The sizes of the surveillance populations were re-estimated for each specific sub-group of the population using the most recent age-specific census estimates on the ratio of men to women, and population level data on HIV prevalence by age group and calendar year using the Thembisa model(24). Since the percentage of IPD cases missing HIV status was high (55% - 68%; **Supplementary Table 1**), we imputed the HIV status of cases with missing data. We performed multiple imputation using chained equations and generated 20 imputed datasets(25, 26). The imputation was based on a logit model using information on sex, age group, IPD status, and carriage. The estimates from the imputed datasets were then combined using Rubin’s rule. We conducted three analyses: 1) an analysis excluding records with missing data, 2) an analysis using imputed values, 3) a sensitivity analysis assigning all cases with missing HIV status to HIV-uninfected. CCRs were recalculated for each record in the dataset and combined using random effects meta-analyses, stratified by sex and HIV status.

The project received ethical approval from London School of Hygiene & Tropical Medicine (Ref: 27394). This activity was reviewed by CDC, deemed not research, and was conducted consistent with applicable federal law and CDC policy (e.g., 45 C.F.R. part 46.102(l)(2), 21 C.F.R. part 56; 42 U.S.C. §241(d); 5 U.S.C. §552a; 44 U.S.C. §3501 et seq.) The funder of the study had no role in study design, data collection, data analysis, data interpretation, or writing of the report.

## Results

### Included studies

A total of 4922 titles and abstracts were screened, and 1165 full texts were assessed for eligibility; 732 publications reported either pneumococcal carriage prevalence or IPD incidence in defined populations. A total of 80 publications from 18 countries reported data on carriage prevalence and IPD incidence in populations in the same country, in overlapping calendar years and age groups and therefore contributed to the analysis (**Figure 1**). For the group level analyses by age-group, country, and time, data from 57,962 participants in carriage surveys and 41,238 IPD cases were included (**Table 1**; **Supplementary Table 2**). Of note, the only upper-middle/high-income countries (UM/HICs) contributing data in adults were England and Wales, and South Africa. We excluded data from South Africa from the UM/HIC grouping at all ages because of the significant differences in CCRs when stratified by HIV status and the heterogeneity introduced when South African CCRs were combined with other UM/HICs.

**Table 1.** Data included in the analyses of CCRs by age group, country income group and PCV status.

| Age group | Income group<sup>1</sup> | PCV period | Total carriage isolates | Total carriage study sample size | Total IPD isolates | Total population under surveillance<sup>2</sup> |
| --- | --- | --- | --- | --- | --- | --- |
| <5 years | L/LMIC | pre-PCV | 5,341 | 7,138 | 417 | 765,288 |
|  |  | post-PCV | 6,666 | 14,750 | 246 | 2,938,546 |
|  | UM/HIC | pre-PCV | 1,902 | 2,264 | 2,771 | 6,710,831 |
|  |  | post-PCV | 6,464 | 14,747 | 4,235 | 57,600,000 |
| 5-14 years | L/LMIC | pre-PCV | 1,006 | 2,495 | 66 | 490139 |
|  |  | post-PCV | 1,727 | 4,649 | 59 | 1,675,842 |
|  | UM/HIC | pre-PCV | 465 | 927 | 702 | 9,175,174 |
|  |  | post-PCV | 56 | 291 | 501 | 13,500,000 |
| 15 years and above | L/LMIC | pre-PCV | 504 | 3,575 | 38 | 769,775 |
|  |  | post-PCV | 1,099 | 6,076 | 133 | 3,116,886 |
|  | UM/HIC | pre-PCV | 375 | 243 | 7,564 | 13,000,000 |
|  |  | post-PCV | 29 | 807 | 24,506 | 53,400,000 |
| Total |  |  | 25,634 | 5,7962 | 41,238 | 16,3142,481 |

Abbreviations: IPD: Invasive pneumococcal disease; L/LMIC: Low and lower-middle income countries; PCV: Pneumococcal conjugate vaccine; UM/HIC: Upper-middle and high-income countries.
<sup>1</sup>This excludes South African data as it was inappropriate to combine South African data with other UM/HICs with much lower HIV prevalences due to the heterogeneity introduced in summary CCR estimates.
<sup>2</sup>If surveillance population sizes were not provided in the publication or by authors, but IPD surveillance was reportedly conducted nationwide, the population size was estimated from the most relevant census (i.e., for Belgium, Brazil, Burkina Faso, Cuba, Denmark, France, Iceland, Norway). If the surveillance population size was provided but was not disaggregated by age group or calendar year, it was divided by the proportion of the population in each age group from the most recent national census, and equally across the calendar years (i.e., Malawi, The Gambia).

For the individual level analyses by sex and HIV status, we received individual-level data on the sex of IPD cases and carriage survey participants from six countries: The Gambia, Kenya, Malawi, Mozambique, England and Wales, and South Africa. Only the South African dataset included the HIV status for both IPD cases and carriage survey participants (**Supplementary Table 1**).

### Population level analysis comparing pre- and post-PCV periods

In under-5s, when grouping serotypes into vaccine-types and non-vaccine types (NVTs, i.e., non-PCV20 types), the CCR for PCV7-types declined after PCV introduction (**Table 2**). In under-5s, CCRs for NVTs declined after PCV introduction in UM/HIC from 102 (95%CI 50-209) to 19 (16–22) post-PCV introduction; but not in low/lower-middle income countries (L/LMICs) where NVT CCRs were 152 (95%CI 103-226) pre-PCV and 154 (119–200) post-PCV (**Table 2**). Differences in CCRs pre-vs. post-PCV introduction were only significant for some serotypes. For example, in under-5s in L/LMICs, ST14 was significantly less invasive post-PCV compared with pre-PCV; in UM/HICs, ST9V, 18C & 35B were less invasive post-PCV compared with pre-PCV. In populations over 15 years of age, in L/LMICs VT and NVT CCRs decreased post-PCV introduction; in England and Wales, CCRs for NVTs increased post-vaccine introduction and in serotype specific analyses ST3 was significantly more invasive post-PCV introduction (and post-PPV23 introduction in adults), compared to the pre-vaccine period (**Supplementary Figure 3**).

**Table 2.** Vaccine-type (VT) and non-vaccine type (NVT) CCRs by PCV introduction status, country income group and age group.

| Serotype-groups | Pre-PCV |  |  |  | Post-PCV |  |  |  |
| --- | --- | --- | --- | --- | --- | --- | --- | --- |
|  | L/LMIC |  | UM/HIC excl. SA |  | L/LMIC |  | UM/HIC excl. SA |  |
|  | CCR | 95%CI | CCR | 95%CI | CCR | 95%CI | CCR | 95%CI |
| **<u>Children <5 yrs of age</u>** |  |  |  |  |  |  |  |  |
| NVT | 152.2 | 102.6 - 225.8 | 102.1 | 49.9 - 208.9 | 154.0 | 118.9 - 199.5 | 18.7 | 16.2 - 21.5 |
| PCV7 types | 152.1 | 112.9 - 204.8 | 80.3 | 60.3 - 107.0 | 100.8 | 74.9 - 135.8 | 45.9 | 31.7 - 66.5 |
| PCV10 additional types | 2891.3 | 1990.1 - 4200.6 | 984.2 | 364.5 - 2657.4 | 1261.4 | 759.1 - 2096.2 | 114.6 | 62.4 - 210.4 |
| PCV13 additional types | 112.2 | 91.5 - 137.6 | 42.2 | 22.4 - 79.4 | 85.6 | 38.7 - 189.6 | 72.5 | 53.4 - 98.4 |
| PCV15 additional types | - | - | 158.5 | 58.2 - 432.1 | - | - | 104.7 | 69.2 - 158.3 |
| PCV20 additional types | 323.1 | 146.0 - 714.7 | 45.0 | 11.0 - 184.3 | 238.3 | 156.1 - 363.6 | 30.0 | 20.2 - 44.5 |
| **<u>5-14 yrs of age</u>** |  |  |  |  |  |  |  |  |
| NVT | 80.2 | 11.9 - 541.1 | 2.7 | 0.9 - 8.2 | 85.5 | 35.3 - 207.1 | 9.5 | 6.5 - 14.0 |
| PCV7 types | 77.9 | 25.9 - 234.0 | 13.0 | 8.8 - 19.1 | 85.9 | 34.8 - 211.9 | 21.9 | 9.2 - 51.8 |
| PCV10 additional types | 1668.3 | 818.0 - 3402.5 | 62.6 | 24.8 - 158.1 | 243.3 | 137.2 - 431.6 | 66.2 | 17.6 - 249.2 |
| PCV13 additional types | 55.2 | 26.1 - 116.7 | 4.6 | 3.0 - 7.0 | 43.3 | 5.0 - 373.3 | 22.9 | 9.6 - 54.9 |
| PCV15 additional types | - | - | 34.8 | 0.3 - 4140.6 | - | - | 43.3 | 13.0 - 145.1 |
| PCV20 additional types | 1278.2 | 26.9 - 61000 | 3.7 | 1.4 - 9.9 | 97.5 | 30.9 - 307.3 | 8.0 | 3.8 - 16.8 |
| **<u>15 years and above</u>** |  |  |  |  |  |  |  |  |
| NVT | 395.6 | 156.9 - 997.3 | 46.1 | 18.8 - 113.0 | 70.8 | 45.4 - 110.6 | 153.8 | 110.2 - 214.6 |
| PCV7 types | 357.8 | 125.1 - 1023.3 | 311.8 | 125.6 - 774.1 | 64.9 | 36.9 - 114.2 | 183.6 | 97.1 - 347.0 |
| PCV10 additional types | 1818.1 | 550.6 - 6003.8 | 491.8 | 144.1 - 1678.2 | 379.7 | 198.1 - 728.0 | 1923.2 | 150.4 - 25000.0 |
| PCV13 additional types | 73.8 | 31.3 - 174.1 | 303.6 | 131.4 - 701.5 | 35.2 | 23.2 - 53.4 | 715.8 | 311.9 - 1642.7 |
| PCV15 additional types | - | - | 317.2 | 40.2 - 2503.1 | - | - | 973.3 | 368.3 - 2572.0 |
| PCV20 additional types | - | - | 213.8 | 60.3 - 757.9 | 129.0 | 45.9 - 362.5 | 229.7 | 115.6 - 456.3 |

Serotype groups are exclusive i.e. NVT are non-PCV20 serotypes. Point estimates are not displayed if 95%CI could not be computed due to data sparsity.
Abbreviations: CCR: case-carrier ratio interpreted as e.g. IPD incidence of 152 cases per 100,000 population, per carrier detected in a cross-sectional carriage survey. L/LMIC: low and lower-middle income countries; PCV: pneumococcal conjugate vaccine; UM/HIC: upper-middle and high-income countries.

### Population level analysis comparing L/LMIC and UM/HIC data

In under-5s, in both pre- and post-PCV periods, CCRs tended to be higher in L/LMIC compared to UM/HIC. Post-PCV introduction in under-5s, CCRs were significantly higher in L/LMICs than in UM/HICs for serotypes 1, 10A, 12F, 15A, 15B, 16F, 35B, 7C and 9N. In populations over 15 years of age, CCRs tended to be higher in UM/HICs compared to L/LMICs, significantly so for serotypes 3, 19A and 8 (**Figure 2**).

**Figure 2.**
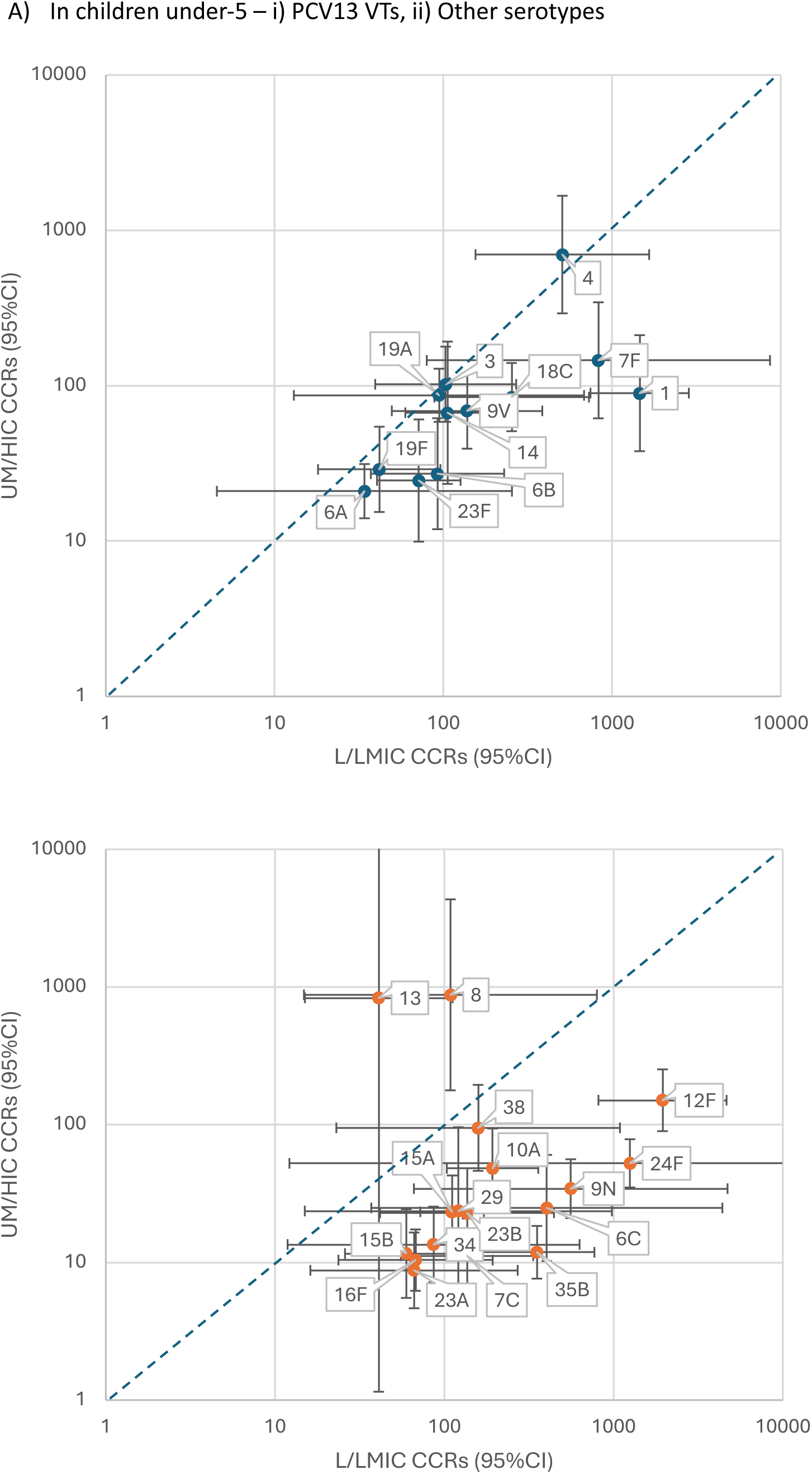
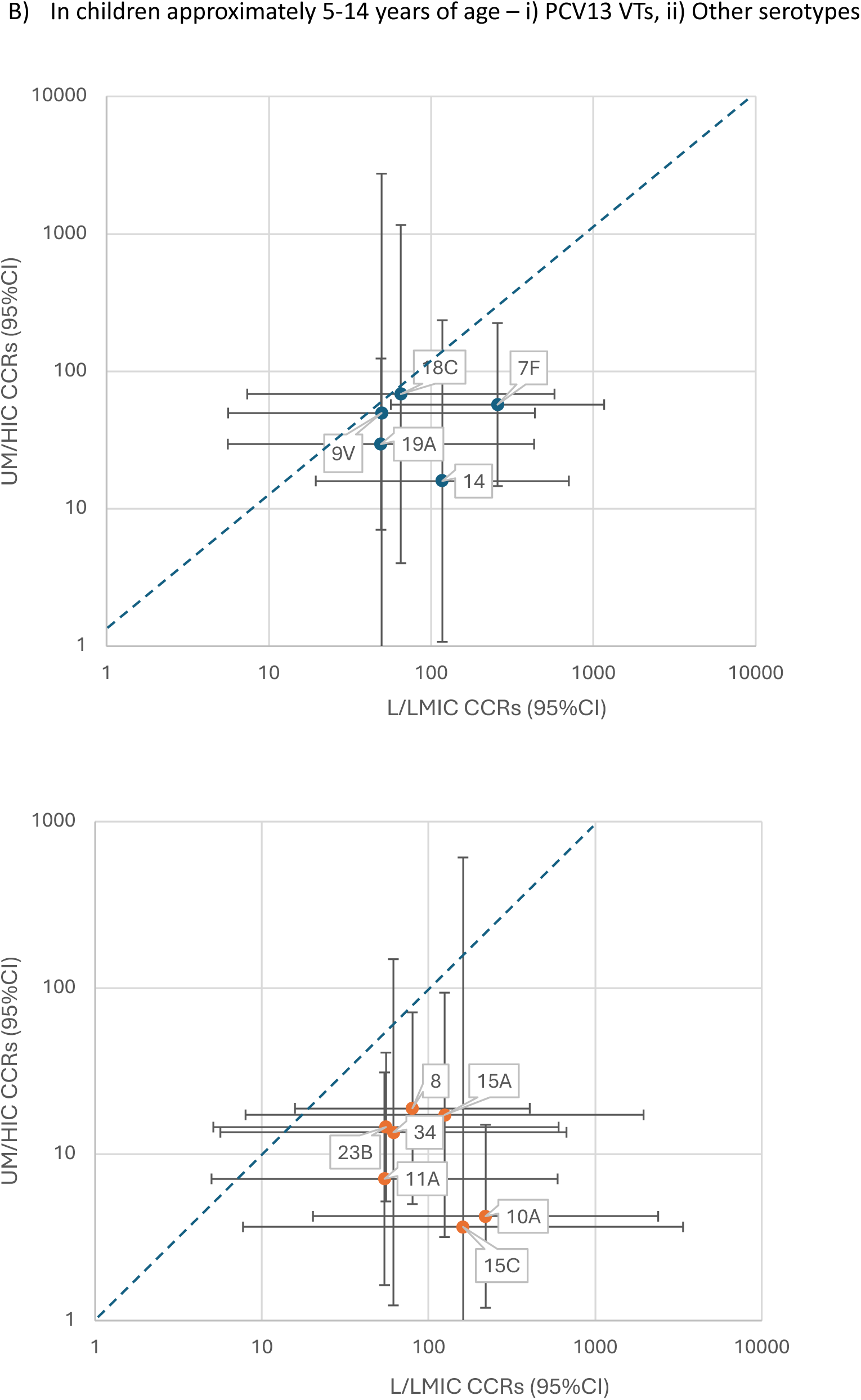
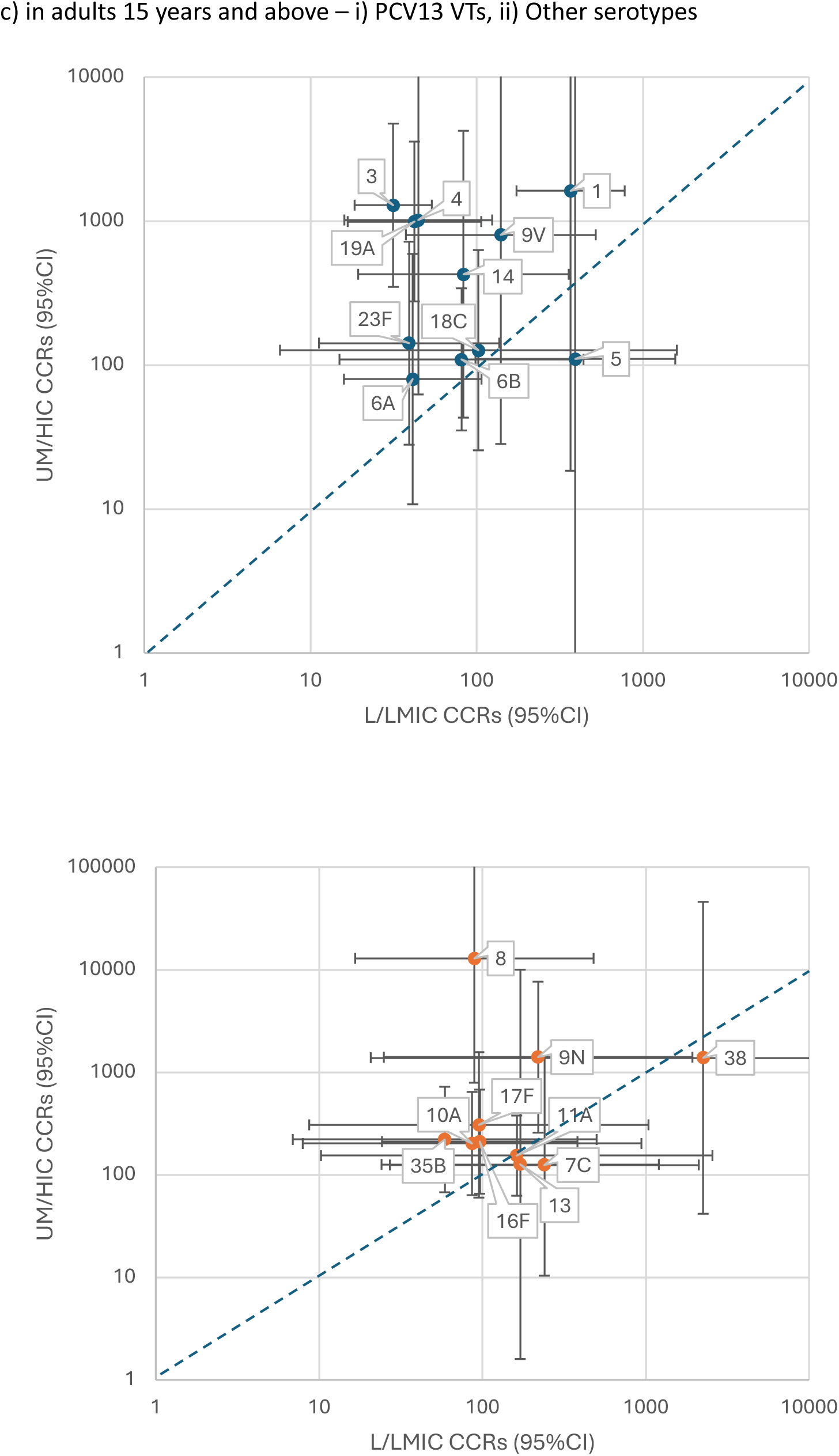
CCRs in UM/HIC compared to L/LMIC, by age group, post-PCV introducAon.

### Population level analysis by age-group

In both pre- and post-PCV periods, CCRs were higher in under-5s and over 15 years-of-age, and lower in 5-14 year olds (**Table 2**; serotype-specific CCRs are available: https://pneumovaxpol.shinyapps.io/Pneumo_CCR_MetaAnalysis/). The 5 additional types included in PCV20 but not in PCV15, are highly invasive in under-5s in L/LMICs and in adults in UM/HICs. In adults in UM/HICs, among a population of carriers of the PCV20 additional serotypes, the expected incidence of IPD would be >200 per 100,000 person years (**Table 2**).

### Population-level trends over time

Where we had data by calendar year prior to and/or following PCV introduction, we analysed whether there were trends over time relative to PCV introduction, restricted to the under-5 age-group. We fitted a linear mixed model to the natural log of the CCRs over years relative to PCV introduction (from −8 to +10 years), controlling clustering by country and serotype. There was significant variation in CCRs over time, but no clear trends in L/LMICs or UM/HICs. There was a downward trend in invasiveness in South African CCRs over time since PCV introduction (**Supplementary Figure 4**).

### Internal calibration

In internal calibration using data from under-5s, the predicted IPD burden was similar to the observed IPD burden for The Gambia, Mozambique and the USA, but was different for Kenya, Malawi and the UK (**Supplementary Table 3)**. Pooled CCRs were weighted to represent datasets which contributed the most IPD cases, and the largest carriage surveys. In an exploratory linear mixed effects model of CCRs from L/LMICs in under-5s, post-PCV, countries with higher care seeking for pneumonia and with a higher proportion of households with handwashing facilities correlated with lower CCRs. Countries with higher prevalences of underweight under-5s were correlated with higher CCRs. The prevalence of HIV in adults, third dose PCV coverage, and the prevalence of biomass fuel use in homes did not correlate with differences in CCRs (**Supplementary Table 4**).

### Individual level analysis by sex and HIV status

There were no significant differences in CCRs by sex; the comparison groups of males and females included a balanced distribution of data from the pre- and post-PCV era (**Supplementary Figure 5**). In South Africa, there were large differences in invasiveness by HIV status across the data pre and post-PCV; all serotypes were more invasive in HIV-infected compared to HIV-uninfected people in the same age stratum (**Figure 3**). The differences in CCRs between HIV-infected and HIV-uninfected groups increased when imputed data on HIV status of IPD cases was used (**Supplementary Figure 6**). Between 2009-2018, the proportion of invasive pneumococcal disease attributable to HIV infection in South Africa was 33.6% (95%CI 30.6-36.6) in children under-5 years of age, 63.8% (95%CI 59.7-67.9) in children 5-14 years, and 76.1% (95%CI 74.8-77.5) in those 15 years and above; however there are likely to be decreasing trends over time (**Supplementary Table 5**).

**Figure 3.**
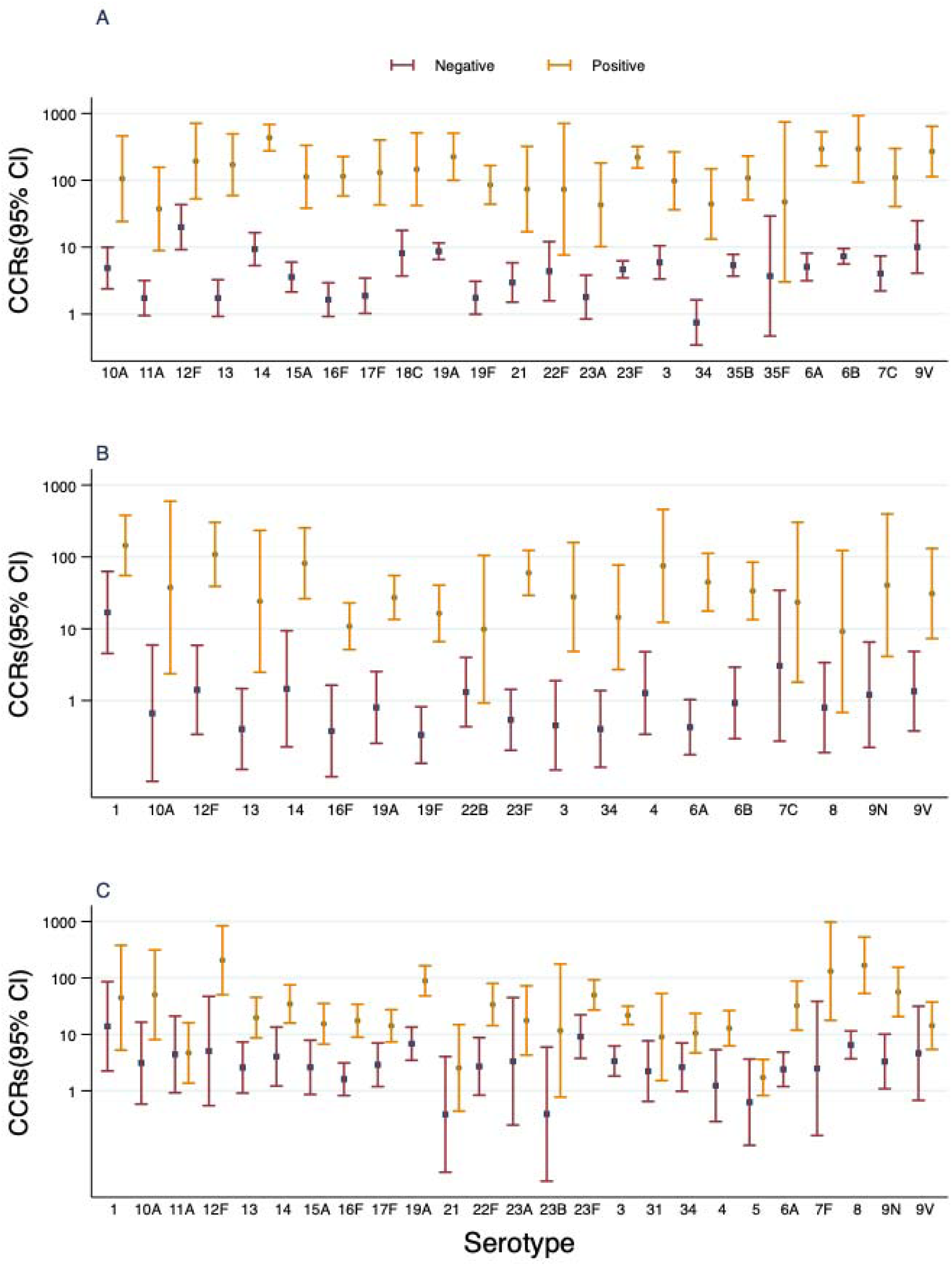
CCRs by HIV status using South African data only, complete-case analysis; A) among children under-5 years of age, B) among children 5-14 years of age, C) among children and adults 15 years and older. ^1^ The percentage of IPD cases missing HIV status was high (55% in children under-5 years of age, 63% in 5-14 year olds, and 68% in 15 year olds and above); the results of the analysis after imputing HIV status for those missing are included in supplementary information.

## Discussion

We aggregated global data from carriage surveys and IPD surveillance. The strength of this review and analysis is its global reach, we have included data from over 40,000 isolates of IPD and over 50,000 carriage survey participants, from a diverse range of settings. We have built on estimates from a prior review(16), which did not disaggregate data by PCV period, and included predominantly high-income country data on children (**Supplementary Table 6**). Our analysis enabled further stratification to refine estimates by country income group, age group and period relative to PCV introduction. We have shown significant differences in invasiveness by serotype, age, population, time period, and no differences by sex. Future statistical or mathematical prediction models of IPD incidence can now use the most relevant of the pooled CCRs for the setting and population of interest.

Among under-5s, the decrease in invasiveness post-PCV introduction for vaccine serotypes is expected as the vaccine confers direct protection against invasion that is independent from the protection from carriage acquisition (27–29). However, there are some surprising differences post-PCV introduction in adults, including a decrease in both NVT and VT CCRs among PCV-unvaccinated adults in L/LMIC. These observations could be explained by reductions in host susceptibility over time and/or that the sensitivity of NVT detection in carriage and diversity of circulating NVTs has increased since PCV introduction, when the dominant vaccine types have been removed(15, 30). This would have overestimated CCRs for NVTs in the pre-vaccine era. In UM/HIC it is notable that we only were able to obtain data on adults from England & Wales and South Africa. This is a significant gap in the literature given the high burden of IPD in adults and the development of vaccines tailored to the prevention of adult IPD (e.g., PCV21)(31).

We estimated that pneumococci are more invasive in L/LMICs, compared to UM/HICs. However, there was substantial variation in CCRs by country. Many factors are likely to influence host susceptibility, including coinfections, malnutrition, air quality, epithelial integrity, and living conditions(32). The comparison of CCRs in L/LMICs and UM/HICs could also be confounded if IPD incidence is, in fact, driven by rates of carriage acquisition, rather than prevalence, and the relationship between carriage prevalence and rates of acquisition differs in L/LMIC compared to UM/HIC (e.g.,with different durations of carriage)(33–36).

In South Africa, serotype-specific CCRs ranged from 2 to 200 times higher in HIV-infected, compared to HIV-uninfected people. Before anti-retroviral therapy (ART) became available, people infected with HIV had 43 times higher incidence rates of IPD than HIV-uninfected populations(37). However, we also showed CCRs in South Africa have been declining over time, which could be explained by the expansion of access to ART, improved diagnostics and prevention of mother to child transmission, and therefore changing levels of immunosuppression among HIV infected populations over time(38, 39). Our comparison of CCRs among HIV-infected and HIV-uninfected did not account for this secular trend as we used all available data from 2010-18.

A limitation of our analysis is the combination of carriage and IPD data from different areas of the same country and different time periods; we attempted to limit the amount of heterogeneity this introduced with our matching criteria. We had to assume that cross sectional carriage prevalence is representative of the population carriage prevalence across an entire year in order to calculate CCRs; however, seasonal variation in carriage has been documented and seasonal variation in invasiveness is likely given that variation in IPD incidence over the course of a year is greater than the variation observed in carriage prevalence(40). In internal validation, pooled CCRs more closely represented datasets which contributed the most IPD cases, which meant they overestimated IPD burden in settings in the same income group, with lower observed IPD incidence (**Supplementary Table 3**). There is substantial variation in invasiveness and IPD burden across different settings that cannot be explained by carriage prevalence and pooled CCRs alone, despite matching data as rigorously as possible, and stratifying the analysis on time period, geography and age group. IPD surveillance sensitivity, including health seeking behaviour, case ascertainment at the facility, sample collection and the rate of laboratory detection of pneumococci, varies across different settings. In children, we may have underestimated the difference in CCRs between L/LMIC and UM/HIC, as IPD surveillance is likely to be of lower sensitivity in L/LMICs. In adults, the reportedly higher CCRs in UM/HIC may be due to low sensitivity of IPD surveillance among adults in L/LMICs, as well as the difference in the age-distribution of the populations (UM/HICs have older adult populations). All the comparisons of pooled CCRs over time and across different settings are confounded by the fact that comparison groups contain different proportions of data from different countries.

Additionally, pneumococci were isolated and serotyped in different ways across the included studies (e.g., by culture and Quellung, latex agglutination and/or mPCR). There were not enough studies using mPCR to compare how the sensitivity of this method of detection/ multiple serotype carriage may have influenced CCRs. Our CCRs are only representative of studies which detected a single serotype. We did not have genomic data on pneumococcal strains; there will be residual confounding in our comparisons of invasiveness across different time periods and countries as it is likely that some of the variability is due to differences in circulating strains of the same serotype(16). However, although sequencing has indicated that strains of serotype 35B in the United States have acquired virulence genes over time(41), in our analysis we do not see an increase in overall invasiveness because carriage prevalence has also increased over time. All the studies included used cross-sectional carriage surveys to estimate carriage prevalence; this will underestimate the prevalence of serotypes with short durations of carriage. To attempt to account for this we imputed very small values of prevalence among studies which reported ‘zero’ prevalence of rarely carried but invasive types.

## Conclusion

High quality IPD surveillance is resource intensive and infeasible for most countries worldwide. We have calculated serotype-specific estimates of pneumococcal invasiveness (available: https://pneumovaxpol.shinyapps.io/Pneumo_CCR_MetaAnalysis/) and have demonstrated invasiveness varies across different populations, age-groups and over time. The most relevant of these pooled estimates could be used in statistical or mathematical models to extrapolate from carriage prevalence data to predict changes in disease incidence where only carriage prevalence is known. This would inform PCV policy decisions including, for example, strategies to increase vaccine coverage, change schedule or introduce a new product.

## Data Availability

All data produced in the present work are contained in the manuscript and associated files.

https://pneumovaxpol.shinyapps.io/Pneumo_CCR_MetaAnalysis/

## Author contributions

Katherine E. Gallagher: accessed data for the formal analysis, project administration, visualization, writing original draft, final responsibility for the decision to submit the manuscript for publication Fredrick Odiwour: accessed data for the formal analysis, writing, review & editing Aisha Adamu, Esther Muthumbi, Eunice W. Kagucia, Laura L Hammitt, Sergio Massora, Betuel Sigaúque, Alberto Chaúque, Leocadia Vilanculos, Jennifer R. Verani, Maria da Gloria Carvalho, Anne von Gottberg, Jackie Kleynhans, Shabir A. Madhi, Courtney P. Olwagen, Grant Mackenzie, Rasheed Salaudeen, Ryan Gierke, Miwako Kobayashi, Stephen Pelton, Inci Yildirim, Stepy Thomas, Amy Tunali, Monica Farley, Todd D. Swarthout, Akuzike Kalizang’oma, Robert S. Heyderman, Neil French, Yoon Choi, Nick Andrews, Shamez Ladhani, Elizabeth Miller: data curation, review & editing C. Bottomley: supervision, review & editing JAG Scott: conceptualization, funding acquisition, supervision, review & editing

## Declaration of interests

None

## Data Sharing

Pooled estimates resulting from the meta-analyses conducted in this paper are available for download via a ShinyApp together with relevant meta-data. Individual level data received from authors for the purposes of this review and metaanalyses, are available from the original investigators only, under applicable data sharing agreements.

## Acknowledgements

Staff time spent conducting the review and meta-analyses was funded by the Wellcome Trust (098532). The funding sources had no role in the study design; collection, analysis and interpretation of data; and in the decision to submit for publication.

We thank the authors of the original research collated here which enabled us to conduct this analysis.

This paper is published with the permission of the Director, Kenya Medical Research Institute. The findings and conclusions in this report are those of the author(s) and do not necessarily represent the official position of the Centers for Disease Control and Prevention.

## Notes

### Competing Interest Statement

The authors have declared no competing interest.

### Author Declarations

The research Ethics committee of the London School of Hygiene & Tropical Medicine gave ethical approval for this work (Ref: 27394). This activity was reviewed by CDC, deemed not research, and waived of the need for ethical approval. The study was conducted consistent with applicable federal law and CDC policy.

### Summary of Updates

a misspelling of an author name

## References

1. Bender RG, Sirota SB, Swetschinski LR, Dominguez R-MV, Novotney A, Wool EE, et al. Global, regional, and national incidence and mortality burden of non-COVID-19 lower respiratory infections and aetiologies, 1990&#x2013;2021: a systematic analysis from the Global Burden of Disease Study 2021. The Lancet Infectious Diseases. 2024;24(9):974–1002.

2. Naghavi M, Mestrovic T, Gray A, Gershberg Hayoon A, Swetschinski LR, Robles Aguilar G, et al. Global burden associated with 85 pathogens in 2019: a systematic analysis for the Global Burden of Disease Study 2019. The Lancet Infectious Diseases. 2024;24(8):868–95.

3. Weiser JN, Ferreira DM, Paton JC. Streptococcus pneumoniae: transmission, colonization and invasion. Nature Reviews Microbiology. 2018;16(6):355–67.

4. Ojal J, Griffiths U, Hammitt LL, Adetifa I, Akech D, Tabu C, et al. Sustaining pneumococcal vaccination after transitioning from Gavi support: a modelling and cost-effectiveness study in Kenya. The Lancet Global Health. 2019;7(5):e644–e54.

5. Rodgers GL, Klugman KP. Surveillance of the impact of pneumococcal conjugate vaccines in developing countries. Human vaccines & immunotherapeutics. 2016;12(2):417–20.

6. Deloria Knoll M, Bennett JC, Garcia Quesada M, Kagucia EW, Peterson ME, Feikin DR, et al. Global Landscape Review of Serotype-Specific Invasive Pneumococcal Disease Surveillance among Countries Using PCV10/13: The Pneumococcal Serotype Replacement and Distribution Estimation (PSERENADE) Project. Microorganisms. 2021;9(4):742.

7. World Health Organization. Pneumococcal vaccines WHO position paper--2019. Releve epidemiologique hebdomadaire / Section d’hygiene du Secretariat de la Societe des Nations = Weekly epidemiological record / Health Section of the Secretariat of the League of Nations. 2019;8(94):85–104.

8. Weinberger DM, Bruden DT, Grant LR, Lipsitch M, O’Brien KL, Pelton SI, et al. Using Pneumococcal Carriage Data to Monitor Postvaccination Changes in Invasive Disease. American Journal of Epidemiology. 2013;178(9):1488–95.

9. Adamu AL, Ojal J, Abubakar IA, Odeyemi KA, Bello MM, Okoromah CAN, et al. The impact of introduction of the 10-valent pneumococcal conjugate vaccine on pneumococcal carriage in Nigeria. Nature communications. 2023;14(1):2666.

10. Choi YH, Andrews N, Miller E. Estimated impact of revising the 13-valent pneumococcal conjugate vaccine schedule from 2+1 to 1+1 in England and Wales: A modelling study. PLoS medicine. 2019;16(7):e1002845.

11. Brueggemann AB, Peto TE, Crook DW, Butler JC, Kristinsson KG, Spratt BG. Temporal and geographic stability of the serogroup-specific invasive disease potential of Streptococcus pneumoniae in children. Journal of Infectious Diseases. 2004;190(7):1203–11.

12. Balsells E, Dagan R, Yildirim I, Gounder PP, Steens A, Muñoz-Almagro C, et al. The relative invasive disease potential of Streptococcus pneumoniae among children after PCV introduction: A systematic review and meta-analysis. J Infect. 2018;77(5):368–78.

13. Flasche S, Hoek AJV, Sheasby E, Waight P, Andrews N, George R. Effect of pneumococcal conjugate vaccination on serotype-specific carriage and invasive disease in England: a cross-sectional study. PLoS medicine. 2011;8.

14. Yildirim I, Hanage WP, Lipsitch M, Shea KM, Stevenson A, Finkelstein J, et al. Serotype specific invasive capacity and persistent reduction in invasive pneumococcal disease. Vaccine. 2010;29(2):283–8.

15. Southern J, Andrews N, Sandu P, Sheppard CL, Waight PA, Fry NK, et al. Pneumococcal carriage in children and their household contacts six years after introduction of the 13-valent pneumococcal conjugate vaccine in England. PloS one. 2018;13(5):e0195799.

16. Løchen A, Truscott JE, Croucher NJ. Analysing pneumococcal invasiveness using Bayesian models of pathogen progression rates. PLOS Computational Biology. 2022;18(2):e1009389.

17. Hammitt LL, Akech DO, Morpeth SC, Karani A, Kihuha N, Nyongesa S. Population effect of 10-valent pneumococcal conjugate vaccine on nasopharyngeal carriage of *Streptococcus pneumoniae* and non-typeable *Haemophilus influenzae* in Kilifi, Kenya: findings from cross-sectional carriage studies. Lancet Glob Heal. 2014;2.

18. Swarthout TD, Fronterre C, Lourenço J, Obolski U, Gori A, Bar-Zeev N, et al. High residual carriage of vaccine-serotype *Streptococcus pneumoniae* after introduction of pneumococcal conjugate vaccine in Malawi. Nature communications. 2020;11(1):2222.

19. Valenciano SJ, Moiane B, Lessa FC, Chaúque A, Massora S, Pimenta FC, et al. Effect of 10-Valent Pneumococcal Conjugate Vaccine on Streptococcus pneumoniae Nasopharyngeal Carriage Among Children Less Than 5 Years Old: 3 Years Post-10-Valent Pneumococcal Conjugate Vaccine Introduction in Mozambique. Journal of the Pediatric Infectious Diseases Society. 2021;10(4):448–56.

20. International Vaccine Access Center (IVAC). VIEW-hub Report: Global Vaccine Introduction and Implementation. Johns Hopkins Bloomberg School of Public Health; 2022

21. Oehlert GW. A Note on the Delta Method. The American Statistician. 1992;46(1):27–9.

22. Hardin JW, Hilbe JM. Estimation and Testing of Binomial and Beta-Binomial Regression Models with and without Zero Inflation. The Stata Journal. 2014;14(2):292–303.

23. Dettori JR, Norvell DC, Chapman JR. Fixed-Effect vs Random-Effects Models for Meta-Analysis: 3 Points to Consider. Global Spine J. 2022;12(7):1624–6.

24. Johnson LF, Meyer-Rath G, Dorrington RE, Puren A, Seathlodi T, Zuma K, et al. The Effect of HIV Programs in South Africa on National HIV Incidence Trends, 2000-2019. Journal of acquired immune deficiency syndromes (1999). 2022;90(2):115–23.

25. Schafer JL. Multiple imputation: a primer. Statistical methods in medical research. 1999;8(1):3–15.

26. Lee JH, Huber Jr J, editors. Multiple imputation with large proportions of missing data: How much is too much? United Kingdom stata users’ group meetings 2011; 2011: Stata Users Group.

27. Black S, Shinefield H. Safety and efficacy of the seven-valent pneumococcal conjugate vaccine: evidence from Northern California. Eur J Pediatr. 2002;161 Suppl 2:S127–31.

28. Hammitt LL, Ojal J, Bashraheil M, Morpeth SC, Karani A, Habib A. Immunogenicity, impact on carriage and reactogenicity of 10-valent pneumococcal non-typeable Haemophilus influenzae protein D conjugate vaccine in Kenyan children aged 1–4 years: a randomized controlled trial. PloS one. 2014;9.

29. Temple B, Tran HP, Dai VTT, Smith-Vaughan H, Licciardi PV, Satzke C, et al. Efficacy against pneumococcal carriage and the immunogenicity of reduced-dose (011+111 and 111+111) PCV10 and PCV13 schedules in Ho Chi Minh City, Viet Nam: a parallel, single-blind, randomised controlled trial. The Lancet Infectious diseases. 2023;23(8):933-44.

30. Olwagen CP, Adrian PV, Madhi SA. Comparison of traditional culture and molecular qPCR for detection of simultaneous carriage of multiple pneumococcal serotypes in African children. Scientific reports. 2017;7(1):4628-.

31. Kobayashi M, Leidner AJ, Gierke R, Farrar JL, Morgan RL, Campos-Outcalt D, et al. Use of 21-Valent Pneumococcal Conjugate Vaccine Among U.S. Adults: Recommendations of the Advisory Committee on Immunization Practices - United States, 2024. MMWR Morb Mortal Wkly Rep. 2024;73(36):793–8.

32. Glennie SJ, Williams NA, Heyderman RS. Mucosal immunity in resource-limited setting: is the battle ground different? Trends Microbiol. 2010;18(11):487–93.

33. Dube FS, Ramjith J, Gardner-Lubbe S, Nduru P, Robberts FJL, Wolter N, et al. Longitudinal characterization of nasopharyngeal colonization with Streptococcus pneumoniae in a South African birth cohort post 13-valent pneumococcal conjugate vaccine implementation. Scientific reports. 2018;8(1):12497.

34. Murad C, Dunne EM, Sudigdoadi S, Fadlyana E, Tarigan R, Pell CL, et al. Pneumococcal carriage, density, and co-colonization dynamics: a longitudinal study in Indonesian infants. International journal of infectious diseases : IJID : official publication of the International Society for Infectious Diseases. 2019.

35. Chaguza C, Senghore M, Bojang E, Lo SW, Ebruke C, Gladstone RA, et al. Carriage Dynamics of Pneumococcal Serotypes in Naturally Colonized Infants in a Rural African Setting During the First Year of Life. Front Pediatr. 2020;8:587730.

36. Metcalf BJ, Waldetoft KW, Beall BW, Brown SP. Variation in pneumococcal invasiveness metrics is driven by serotype carriage duration and initial risk of disease. Epidemics. 2023;45:100731.

37. Meiring S, Cohen C, Quan V, de Gouveia L, Feldman C, Karstaedt A, et al. HIV Infection and the Epidemiology of Invasive Pneumococcal Disease (IPD) in South African Adults and Older Children Prior to the Introduction of a Pneumococcal Conjugate Vaccine (PCV). PloS one. 2016;11(2):e0149104.

38. Rohr JK, Manne-Goehler J, Gómez-Olivé FX, Kahn K, Bärnighausen TW. The HIV Care Cascade for Older Adults in Rural South Africa: A Longitudinal Cohort Study (2014-2019). Journal of acquired immune deficiency syndromes (1999). 2024;96(4):334–40.

39. Lippman SA, El Ayadi AM, Grignon JS, Puren A, Liegler T, Venter WDF, et al. Improvements in the South African HIV care cascade: findings on 90-90-90 targets from successive population-representative surveys in North West Province. J Int AIDS Soc. 2019;22(6):e25295.

40. Domenech de Cellès M, Arduin H, Lévy-Bruhl D, Georges S, Souty C, Guillemot D, et al. Unraveling the seasonal epidemiology of pneumococcus. Proceedings of the National Academy of Sciences. 2019;116(5):1802.

41. Chochua S, Metcalf B, Li Z, Walker H, Tran T, McGee L, et al. Invasive Serotype 35B Pneumococci Including an Expanding Serotype Switch Lineage, United States, 2015–2016. Emerging Infectious Disease journal. 2017;23(6):922.

