## Supplementary for "Serotype-specific pneumococcal invasiveness: a global meta-analysis of paired estimates of disease incidence and carriage prevalence"

### Supplementary files

Authors: Katherine E. Gallagher[1,2*] & Fredrick Odiwour[1*], Christian Bottomley[2], John Ojal[1], Aisha Adamu[3], Esther Muthumbi[1], Eunice W. Kagucia[1], Laura L Hammitt[4], Sergio Massora[5], Betuel Sigaúque[5], Alberto Chaúque[5], Leocadia Vilanculos[5], Jennifer R. Verani[6], Maria da Gloria Carvalho[6], Anne von Gottberg[7,8], Jackie Kleynhans[7,9], Shabir A. Madhi[10], Courtney P. Olwagen[10], Grant Mackenzie[11], Rasheed Salaudeen[11], Ryan Gierke[6], Miwako Kobayashi[6], Stephen Pelton[12], Inci Yildirim[12-16], Stepy Thomas[17], Amy Tunali[17], Monica Farley[17], Todd D. Swarthout[18,19], Akuzike Kalizang'oma[18,19], Robert S. Heyderman [18,19], Neil French [19,20], Yoon Choi[21], Nick Andrews[21], Shamez Ladhani[21], Elizabeth Miller[2], J. Anthony G. Scott [1,2]

[1] Epidemiology & Demography Department, KEMRI-Wellcome Trust Research Programme (KWTRP), Kilifi, Kenya
[2]London School of Hygiene & Tropical Medicine (LSHTM), UK
[3]International Health and Tropical Medicine, Centre for Tropical Medicine and Global Health, Nuffield Department of Medicine, University of Oxford, Oxford, United Kingdom.
[4]International Vaccine Access Center, Johns Hopkins Bloomberg School of Public Health, Baltimore, MD, USA
[5]Centro de Investigaçao em Saúde de Manhiça (CISM), Mozambique
[6]National Center for Immunization and Respiratory Diseases, Centers for Disease Control and Prevention, Atlanta, USA
[7]National Institute for Communicable Diseases of the National Health Laboratory Service, Johannesburg, South Africa
[8]Department of Clinical Microbiology and Infectious Diseases (CMID), School of Pathology, Faculty of Health Sciences, University of the Witwatersrand, Johannesburg, South Africa
[9]School of Public Health, Faculty of Health Sciences, University of the Witwatersrand, Johannesburg, South Africa
[10] South African Medical Research Council Vaccines and Infectious Diseases Analytics Research Unit, Faculty of Health Sciences, University of the Witwatersrand, Johannesburg, South Africa
[11]The Medical Research Council Unit The Gambia at the London School of Hygiene & Tropical Medicine
[12]Boston University School of Medicine and Public Health, USA
[13]Department of Pediatrics, Section of Infectious Diseases and Global Health; Yale University School of Medicine, New Haven, CT
[14]Department of Epidemiology of Microbial Diseases, Yale School of Public Health, New Haven, CT
[15]Yale Institute for Global Health, Yale University, New Haven, CT
[16]Yale Center for Infection and Immunity, Yale University, New Haven, CT
[17] Georgia Emerging Infections Program, Emory University School of Medicine, USA
[18]Research Department of Infection, Division of Infection and Immunity, University College London
[19]Malawi-Liverpool-Wellcome Clinical Research Programme, Malawi
[20]Institute of Infection, Veterinary and Ecological Sciences, University of Liverpool,
[21]UK Health Security Agency (UKHSA), London, England.

**Supplementary Table 1. Observed data included in the individual level analyses by HIV status and by sex[1]**

**A. Data included in the analyses of CCRs by HIV status**

| Country | IPD surveillance area | Carriage survey area | Matched survey years | Matched ages | HIV status | IPD surveillance population size | Carriage sample size |
|---|---|---|---|---|---|---|---|
| **South Africa** | Nationwide | Agincourt, Mpumalanga province, rural NE South Africa | 2009-15, 2017-18 | All ages (<5, 5-14, 15+ years) | Negative | 414003922 | 14264 |
| | | | | | Positive | 58491516 | 4332 |

**B. Data included in the analyses of CCRs by sex**

| Country | IPD surveillance area | Carriage survey area | Matched survey years | Matched ages | Sex | IPD surveillance population size | Carriage sample size |
|---|---|---|---|---|---|---|---|
| **Gambia** | Basse HDSS | Basse HDSS | 2009, 2015-17 | All ages (<5, 5-14, 15+ years) | Female | 366206 | 4979 |
| | | | | | Male | 360754 | 4090 |
| **Kenya** | Kilifi HDSS | Kilifi HDSS | 2004, 2006-10, 2012-19 | All ages (<5 ,5-14, 15+ years) | Female | 1788644 | 6956 |
| | | | | | Male | 1591988 | 5330 |
| **Malawi** | Blantyre | Blantyre | 2015-2018 | All ages (<5, 5-14, 15+ years) | Female | 1635546 | 3915 |
| | | | | | Male | 1527985 | 3297 |
| **Mozambique** | Manhiça | Manhiça | 2012-16, 2018-19, 2021-22 | <5 years | Female | 115092 | 2223 |
| | | | | | Male | 111011 | 2234 |
| **England & Wales** | Nationwide | Hertfordshire, Gloucestershire | 2001-02, 2008-09, 2012-13, 2015-16, 2018 | All ages (<5,5-14,15+ years) | Female | 40038218 | 1391 |
| | | | | | Male | 36310574 | 1015 |
| **South Africa** | Nationwide | Agincourt, Mpumalanga province, rural northeast South Africa | 2009-15, 2017-18 | All ages (<5,5-14,15+ years) | Female | 220007324 | 11247 |
| | | | | | Male | 167261773 | 4240 |

Abbreviations: HDSS: Health and Demographic Surveillance Site; HIV: human immunodeficiency virus; IPD: Invasive Pneumococcal Disease.

[1]Individual level data from the USA was provided by authors but it was not disaggregated by sex or HIV status.

C. **The proportion of IPD cases with missing HIV status by age-group, year, and sex in South Africa matched data**

| Characteristic | HIV-uninfected | | HIV-infected | | HIV missing | |
|---|---|---|---|---|---|---|
| | n | (%) | n | (%) | n | (%) |
| **Age-group** | | | | | | |
| <5 years | 1202 | (29.3) | 660 | (16.1) | 2239 | (54.6) |
| 5-14 years | 192 | (13.2) | 350 | (24.0) | 918 | (62.9) |
| >=15 years | 931 | (6.2) | 3824 | (25.3) | 10335 | (68.5) |
| Missing | 0 | | 3 | (0.5) | 590 | (99.5) |
| **Year** | | | | | | |
| 2009 | 315 | (9.3) | 821 | (24.2) | 2253 | (66.5) |
| 2010 | 302 | (9.2) | 857 | (26.2) | 2109 | (64.5) |
| 2011 | 322 | (11.7) | 683 | (24.9) | 1739 | (63.4) |
| 2012 | 278 | (11.6) | 548 | (22.9) | 1569 | (65.5) |
| 2013 | 291 | (13.5) | 447 | (20.7) | 1425 | (65.9) |
| 2014 | 277 | (13.8) | 393 | (19.6) | 1336 | (66.6) |
| 2015 | 187 | (10.0) | 393 | (21.0) | 1293 | (69.0) |
| 2017 | 212 | (12.0) | 348 | (19.7) | 1210 | (68.4) |
| 2018 | 141 | (8.6) | 347 | (21.2) | 1148 | (70.2) |
| **Sex** | | | | | | |
| Females | 1024 | (9.2) | 2689 | (25.0) | 7047 | (65.5) |
| Males | 1300 | (12.8) | 2144 | (21.1) | 6735 | (66.2) |
| Missing | 1 | (0.3) | 4 | (1.3) | 300 | (98.9) |

**D. The proportion of carriage survey participants with missing HIV status by age-group, year, and sex in South Africa matched data**

| Characteristic | HIV-uninfected | | HIV-infected | | HIV missing | |
|---|---|---|---|---|---|---|
| | n | (%) | n | (%) | n | (%) |
| **Age-group** | | | | | | |
| <5 years | 8216 | (81.5) | 1642 | (16.3) | 218 | (2.2) |
| 5-14 years | 1285 | (69.4) | 411 | (22.2) | 157 | (8.5) |
| >=15 years | 4763 | (59.5) | 2279 | (28.5) | 959 | (12.0) |
| Missing | | | | | | |
| **Year** | | | | | | |
| 2009 | 1607 | (68.6) | 307 | (13.1) | 430 | (18.3) |
| 2010 | 2421 | (60.9) | 1370 | (34.5) | 182 | (4.6) |
| 2011 | 3145 | (75.6) | 509 | (12.2) | 508 | (12.2) |
| 2012 | 1403 | (54.6) | 1166 | (45.4) | 1 | (0.0) |
| 2013 | 2064 | (82.1) | 250 | (10.0) | 199 | (7.9) |
| 2014 | 652 | (81.8) | 128 | (16.1) | 17 | (2.1) |
| 2015 | 299 | (55.3) | 240 | (44.4) | 2 | (0.4) |
| 2017 | 357 | (75.2) | 118 | (24.8) | 0 | (0.0) |
| 2018 | 2316 | (90.1) | 244 | (9.5) | 12 | (0.5) |
| **Sex** | | | | | | |
| Females | 7831 | (69.5) | 2616 | (23.2) | 817 | (7.3) |
| Males | 3567 | (18.1) | 358 | (8.4) | 315 | (7.4) |
| Missing | 2866 | (64.5) | 1358 | (30.6) | 219 | (4.9) |

**Supplementary Table 2. All available matched IPD and carriage data, by country**

| Country | IPD surveillance area | IPD surveillance years | IPD surveillance ages | Carriage area | Carriage survey years | Carriage ages | Pneumococcal vaccine programme[1] |
|---|---|---|---|---|---|---|---|
| **HIC** | | | | | | | |
| **Belgium (1-3)** | Nationwide | 2015-2018 | 0-30 months | Flanders, Wallonia, Brussels | 2016-2018 | 0-30 months | Child PCV13 |
| | | 2017-2018 | <5 years | Flanders, Wallonia, Brussels | 2016-2017 | 6 months-≤30 months | Child PCV13 |
| **Denmark (4, 5)** | Nationwide | 2000-2007 | 2-4 years | Roskilde | 1999-2000 | 12-72 months | None |
| **England & Wales (6-15)** | Nationwide | 2001-2002 | All (incl. sub-groups of <5, 5-14, ≥15 yrs) | Hertfordshire, Gloucestershire | 2001-2002 | All (incl. sub-groups of <5, 5-14, ≥15 yrs) | None |
| | | 2008-2009 | All (incl. sub-groups of <5, 5-14, ≥15 yrs) | | 2008-2009 | All (incl. sub-groups of <5, 5-14, ≥15 yrs) | Child PCV7, Adult PPV23 |
| | | 2012-13, 2015-16, 2018 | All (incl. sub-groups of <5, 5-14, ≥15 yrs) | | 2012-13, 2015-16, 2018 | All (incl. sub-groups of <5, 5-14, ≥15 yrs) | Child PCV13, Adult PPV23 |
| **France (16-19)** | Nationwide | 2001-2002 | <2 years | Alpes Maritimes | 2002 | 3-40 months | None |
| | | 2001-2002 | <2 years | Alpes Maritimes | 2002 | 3-40 months | None |
| | | 2006 | <2 years | Alpes Maritimes | 2006 | 3-40 months | Child PCV13 |
| | | 2008-2009 | <2 years | Alpes Maritimes | 2008 | 3-40 months | Child PCV13 |
| | | 2012 | <2 years | Alpes Maritimes | 2012 | 3-40 months | Child PCV13 |
| **Greenland (20, 21)** | Nationwide | 2010-2020 | All ages | Tasiilaq, Sisimiut | 2011 | 0-6 years | Child PCV13 |
| | Nationwide | 2010-2020 | All ages | Tasiilaq, Sisimiut | 2013 | 0-6 years | Child PCV13 |
| **Iceland (22, 23)** | Nationwide | 2007-2011 | <7 years | Nationwide | 2010 | <7 years | None |
| | Nationwide | 2007-2011 | <7 years | Nationwide | 2009 | <7 years | None |
| | Nationwide | 2007-2011 | <7 years | Nationwide | 2011 | <7 years | None |
| **Norway (24-26)** | Nationwide | 2006 | <5 years | Oslo | 2006 | 0-6 years | Child PCV13 |
| | Nationwide | 2006 | <5 years | Oslo | 2006 | 0-6 years | Child PCV13 |
| | Nationwide | 2008 | <5 years | Oslo | 2008 | 0-6 years | Child PCV13 |
| **Portugal (27-32)** | Nationwide | 2008-2012 | <18 yrs | Coimbra | 2008 | 3 months to <7 years | None |
| | Nationwide | 2008-2012 | <18 yrs | Oeiras (urban) | 2010-2011 | ≤6 years | None |

| | | | | | | | |
|---|---|---|---|---|---|---|---|
| | Nationwide | 2008-2012 | <18 yrs | Lisbon district | 2010 | 3 months to <7 years | None |
| | Nationwide | 2008-2012 | <18 yrs | Montemor-o-Novo (rural) | 2010-2011 | ≤6 years | None |
| | Nationwide | 2008-2012 | <18 yrs | Coimbra | 2009 | 3 months to <7 years | None |
| | Nationwide | 2008-2012 | <18 yrs | Lisbon district | 2009 | 3 months to <7 years | None |
| | Nationwide | 2008-2012 | <18 yrs | Montemor-o-Novo (rural) | 2009-2010 | ≤6 years | None |
| | Nationwide | 2008-2012 | <18 yrs | Oeiras (urban) | 2009-2010 | ≤6 years | None |
| | Nationwide | 2012-2015 | <18 yrs | Oeiras (urban) | 2011-2012 | <6 years | None |
| | Nationwide | 2012-2015 | <18 yrs | Oeiras (urban) | 2015-2016 | <6 years | None |
| | Nationwide | 2012-2015 | <18 yrs | Montemor-o-Novo (rural) | 2015-2016 | <6 years | None |
| | Nationwide | 2012-2015 | <18 yrs | Montemor-o-Novo (rural) | 2011-2012 | <6 years | None |
| | Nationwide | 2015-2018 | <18 yrs | Oeiras (urban) | 2015-2016 | <6 years | Child PCV13 |
| | Nationwide | 2015-2018 | <18 yrs | Montemor-o-Novo (rural) | 2015-2016 | <6 years | Child PCV13 |
| **USA (33-38)** | National | 2010-17 | <5 years | Georgia | 2010-17 | <5 years | Child PCV13, Adult PPV23 |
| **Czech Republic (39, 40)** | Surveillance network of 55% of popn | 2000-2006 | 1-4 years | 9 cities | 2004-2005 | 3-5 years | None |
| **UMIC** | | | | | | | |
| **Brazil (41, 42)** | Nationwide | 2011 | <5 years | Goiania | 2010-2011 | 7-18 months | Child PCV10 |
| **Cuba (42, 43)** | Nationwide | 2013 | <5 years | Cienfuegos | 2013 | 2-18 months | None |
| **South Africa (44-51)** | Nationwide | 2010 | All (incl. sub-groups of <5, 5-14, ≥15 yrs) | Agincourt, Mpumalanga province | 2010 | All (incl. sub-groups of <5, 5-14, ≥15 yrs) | Child PCV7, Adult PPV23 |
| | Nationwide | 2011-2018 | All (incl. sub-groups of <5, 5-14, ≥15 yrs) | Agincourt, Mpumalanga province | 2011-18 | All (incl. sub-groups of <5, 5-14, ≥15 yrs) | Child PCV13, Adult PPV23 |
| **LMIC** | | | | | | | |
| **Kenya (52-54)** | Kilifi HDSS | 2004, 2006-10 | All (incl. sub-groups of <5, 5-14, ≥15 yrs) | Kilifi HDSS | 2004, 2006-10 | All (incl. sub-groups of <5, 5-14, ≥15 yrs) | None |

| | | | | | | | |
|---|---|---|---|---|---|---|---|
| | Kilifi HDSS | 2012-19 | All (incl. sub-groups of <5, 5-14, ≥15 yrs) | Kilifi HDSS | 2012-19 | All (incl. sub-groups of <5, 5-14, ≥15 yrs) | Child PCV10 |
| **LIC** | | | | | | | |
| **Burkina Faso (55, 56)** | Nationwide | 2015 | <1 years | Bobo-Dioulasso | 2015 | <5 years | Child PCV13 |
| | Nationwide | 2017 | <1 years | Bobo-Dioulasso | 2017 | <5 years | Child PCV13 |
| **Gambia (57-70)** | Basse HDSS | 2009 | All (incl. sub-groups of <5, 5-14, ≥15 yrs) | Basse HDSS | 2009 | All (incl. sub-groups of <5, 5-14, ≥15 yrs) | None |
| | Basse HDSS | 2015-17 | All (incl. sub-groups of <5, 5-14, ≥15 yrs) | Basse HDSS | 2015-17 | All (incl. sub-groups of <5, 5-14, ≥15 yrs) | Child PCV13 |
| **Malawi (71-74)** | Blantyre | 2015-2018 | All (incl. sub-groups of <5, 5-14, ≥15 yrs) | Blantyre | 2015-2019 | All (incl. sub-groups of <5, 5-14, ≥15 yrs) | Child PCV13 |
| | Blantyre | 2018 | All (incl. sub-groups of <5, 5-14, ≥15 yrs) | Blantyre | 2018 | All (incl. sub-groups of <5, 5-14, ≥15 yrs) | Child PCV13 |
| **Mozambique (75-80)** | Manhiça | 2012 | <5 years | Manhiça | 2012 | <5 years | None |
| | Manhiça | 2013-14, 2019, 2021-22 | <5 years | Manhiça | 2013-14, 2019, 2021-22 | <5 years | Child PCV10 |

Abbreviations: HDSS: Health and Demographic Surveillance Site; HIC: High-Income Country; HIV: human immunodeficiency virus; IPD: Invasive Pneumococcal Disease; LIC: low-income country; LMIC: lower-middle income country; UMIC: upper-middle income country.

[1] The vaccine programme in the population included in the study is listed, 'Child' indicates the vaccine listed was targeted at children in the period included in the review; 'Adult' indicates the vaccine listed was targeted at adults (generally adults over 65 years of age) in the period and population listed as included in the review.

**Supplementary Table 3. Internal validation of CCRs using datasets shared by authors which are well matched in terms of age, calendar year, and place**

**A) Summary of internal validation – VT and NVT[1] - using ST-specific CCRs**

| Data | Serogroups | Observed carriage | % | Observed number of IPD isolates | Observed IPD incidence/ 100,000 pyrs | Predicted IPD incidence/ 100,000 pyrs (95% CI) |
|---|---|---|---|---|---|---|
| **L/LMIC** | | | | | | |
| **Kenya (2012-19) under-5s** | PCV10 types | 109/1424 | **7.7%** | 12 | 3.61 | 10.2 (4.47-24.3) |
| | NVT | 781/1424 | **54.8%** | 25 | 7.53 | 62.4 (21.4-291) |
| **Malawi (2015-18) under-5s** | PCV13 types | 630/3583 | **17.6%** | 31 | 6.16 | 22.8 (9.63-66.5) |
| | NVT | 836/3583 | **23.3%** | 2 | 0.40 | 21.1 (5.30-135.6) |
| **The Gambia (2015-17) under-5s** | PCV13 types | 293/1925 | **15.2%** | 11 | 12.9 | 16.0 (5.75-63.6) |
| | NVT | 1042/1925 | **54.1%** | 45 | 52.6 | 53.9 (17.1-247.7) |
| **Mozambique (2013-23) under-5s** | PCV10 types | 604/3959 | **14.7%** | 23 | 11.9 | 16.0 (7.14 – 37.6) |
| | NVT | 1845/3959 | **41.0%** | 19 | 9.08 | 57.3 (19.1-273) |
| **UM/HIC** | | | | | | |
| **England & Wales (2012-18) under-5s** | PCV13 types | 5/649 | **0.8%** | 225 | 8.67 | 0.63 (0.38-1.03) |
| | NVTs | 303/649 | **46.7%** | 978 | 37.6 | 11.6 (6.56-20.7) |
| **USA (2010-17), under-5s** | PCV13 types | 126/5079 | **2.5%** | 506 | 3.00 | 1.95 (1.25-3.09) |
| | NVT | 1341/5079 | **26.4%** | 1077 | 6.38 | 5.04 (2.77-9.48) |

[1] NVT are defined differently in each country depending on the listed PCV in use.

In LIC/LMIC: Multiplying observed carriage proportions with the available pooled CCRs, resulted in overestimates of the IPD burden in Kenya, Malawi, but an accurate prediction of overall burden in The Gambia and Mozambique. The variance around the CCRs was determined by the number of IPD cases detected, the number of carriers and the size of the carriage surveys; this variance was then used to assign weights to CCRs when they were pooled across settings. The Gambia reported much higher numbers of IPD isolates than other L/LMIC settings and therefore the CCRs were heavily weighted to represent this setting.

In UM/HIC: good prediction in USA, poor prediction in UK. A higher number of IPD isolates and larger carriage dataset was available from USA which will have weighted CCRs more heavily towards representing USA data.

The weighting will differ by serotype; however, as the CCRs are applied to each serotype individually and then the predicted IPD summed across serotypes. The representativeness of the predicted burden will be influenced by the serotype distribution of the contributing data to the CCRs, and the sensitivity of the observed serotype-specific carriage prevalence.

B) **Summary of internal validation – VT and NVT – using serogroup CCRs**

| **Serogroups** | **Observed carriage** | | | **Observed IPD** | | | **Predicted IPD** | |
|---|---|---|---|---|---|---|---|---|
| | **n** | **N** | **%** | **Number of isolates** | **Population under surveillance** | **Incidence (/100,000 pyrs)** | **Incidence (/100,000 pyrs)** | **95%CI** |
| **England & Wales, 2012-18, under-5s (National IPD data, carriage data from Herefordshire, Gloucestershire)** | | | | | | | | |
| NVT | 215 | 649 | 33.1% | 499 | 2599309 | 19.20 | 6.19 | 5.37-7.12 |
| PCV7 VT | 1 | 649 | 0.2% | 35 | 2599309 | 1.35 | 0.07 | 0.05-0.10 |
| add. PCV10 VT | 0 | 649 | 0.0% | 58 | 2599309 | 2.23 | 0.00 | 0-0 |
| add. PCV13 VT | 4 | 649 | 0.6% | 132 | 2599309 | 5.08 | 0.45 | 0.33-0.61 |
| add. PCV15 VT | 22 | 649 | 3.4% | 148 | 2599309 | 5.69 | 3.55 | 2.35-5.37 |
| add. PCV20 VT | 66 | 649 | 10.2% | 331 | 2599309 | 12.73 | 3.05 | 2.05-4.53 |
| **USA, 2010-17, under-5s (Sub-national IPD data, carriage data from Georgia)** | | | | | | | | |
| NVT | 1029 | 5079 | 20.3% | 591 | 16881820 | 3.50 | 3.79 | 3.28-4.36 |
| PCV7 VT | 22 | 5079 | 0.4% | 62 | 16881820 | 0.37 | 0.20 | 0.14-0.29 |
| add. PCV10 VT | 0 | 5079 | 0.0% | 81 | 16881820 | 0.48 | 0.00 | |
| add. PCV13 VT | 104 | 5079 | 2.0% | 363 | 16881820 | 2.15 | 1.48 | 1.09-2.01 |
| add. PCV15 VT | 52 | 5079 | 1.0% | 280 | 16881820 | 1.66 | 1.07 | 0.71-1.62 |
| add. PCV20 VT | 260 | 5079 | 5.1% | 206 | 16881820 | 1.22 | 1.54 | 1.03-2.28 |
| **Malawi, 2015-18, under-5s** | | | | | | | | |
| NVT | 747 | 3583 | 20.8% | 2 | 503001 | 0.40 | 32.11 | 24.79-41.59 |
| PCV7 VT | 379 | 3583 | 10.6% | 12 | 503001 | 2.39 | 10.66 | 7.92-14.36 |
| add. PCV10 VT | 23 | 3583 | 0.6% | 14 | 503001 | 2.78 | 8.10 | 4.87-13.46 |
| add. PCV13 VT | 228 | 3583 | 6.4% | 5 | 503001 | 0.99 | 5.45 | 2.46-12.06 |
| add. PCV15 VT | 5 | 3583 | 0.1% | 0 | 503001 | 0.00 | . | |
| add. PCV20 VT | 84 | 3583 | 2.3% | 0 | 503001 | 0.00 | 5.59 | 3.66-8.52 |
| **The Gambia, 2015-17, under-5s** | | | | | | | | |
| NVT | 847 | 1925 | 44.0% | 32 | 85600 | 37.38 | 67.76 | 52.32-87.78 |
| PCV7 VT | 162 | 1925 | 8.4% | 5 | 85600 | 5.84 | 8.48 | 6.30-11.43 |
| add. PCV10 VT | 5 | 1925 | 0.3% | 4 | 85600 | 4.67 | 3.28 | 1.97-5.44 |
| add. PCV13 VT | 126 | 1925 | 6.5% | 2 | 85600 | 2.34 | 5.60 | 2.53-12.41 |
| add. PCV15 VT | 2 | 1925 | 0.1% | 0 | 85600 | 0.00 | . | |
| add. PCV20 VT | 193 | 1925 | 10.0% | 13 | 85600 | 15.19 | 23.89 | 15.65-36.45 |
| **Mozambique, 2013-22, under-5s** | | | | | | | | |
| NVT | 1467 | 3959 | 37.1% | 11 | 209341 | 5.25 | 57.06 | 44.06-73.92 |
| PCV7 VT | 598 | 3959 | 15.1% | 12 | 209341 | 5.73 | 15.23 | 11.31-20.51 |
| add. PCV10 VT | 6 | 3959 | 0.2% | 11 | 209341 | 5.25 | 1.91 | 1.15-3.18 |
| add. PCV15 VT | 9 | 3959 | 0.2% | 0 | 209341 | 0.00 | . | |
| add. PCV20 VT | 369 | 3959 | 9.3% | 8 | 209341 | 3.82 | 22.21 | 14.55-33.89 |
| **Kenya, 2012-19, under-5s** | | | | | | | | |
| NVT | 607 | 1424 | 42.6% | 20 | 332032 | 6.02 | 65.64 | 50.68-85.04 |
| PCV7 VT | 106 | 1424 | 7.4% | 4 | 332032 | 1.20 | 7.50 | 5.58-10.11 |

| | | | | | | | | |
|---|---|---|---|---|---|---|---|---|
| add. PCV10 VT | 3 | 1424 | 0.2% | 8 | 332032 | 2.41 | 2.66 | 1.60-4.42 |
| add. PCV15 VT | 2 | 1424 | 0.1% | 0 | 332032 | 0.00 | . | |
| add. PCV20 VT | 172 | 1424 | 12.1% | 5 | 332032 | 1.51 | 28.78 | 18.85-43.92 |

[1] In LIC/LMIC: good prediction in the Gambia and Mozambique (CCRs from the Gambia and Mozambique will have been weighted towards these regions due to the number of IPD isolates and carriage data contributed for VTs and NVTs), but poor prediction in Kenya and Malawi.

In UM/HIC: good prediction in USA, poor prediction in UK. A higher number of IPD isolates and larger carriage dataset was available from USA which will have weighted CCRs more heavily towards representing USA data.

The weighting will differ by serotype. The representativeness of the predicted burden will be influenced by the serotype distribution of the contributing data to the CCRs, and the sensitivity of the observed serotype-specific carriage prevalence.

**Supplementary Table 4. Linear Mixed effects model of factors associated with variation in CCRs restricted to LMIC, under-5s, post-PCV**

| | Crude analysis | | | Adjusted analysis for care seeking | | |
|---|---|---|---|---|---|---|
| | **Ratio** | **95%CI** | **p-value** | **aRatio** | **95%CI** | **p-value** |
| **HIV prevalence in adults 15-49** | 0.99 | 0.95-1.03 | 0.513 | | | |
| **Children U5 U-weight** | 1.16 | 1.03-1.30 | 0.013 | 1.03 | 0.87-1.21 | 0.74 |
| **Care seeking pneumonia** | 0.95 | 0.92-0.97 | 0.0001 | 0.95 | 0.92-0.99 | 0.001 |
| **Handwashing** | 0.98 | 0.96-0.99 | 0.004 | 0.99 | 0.97-1.00 | 0.14 |
| **PCV3 coverage** | 1.00 | 0.98-1.01 | 0.636 | | | |
| **Biomass use** | 0.99 | 0.96-1.02 | 0.512 | | | |

CCRs included in the analysis from Kenya, Malawi, Mozambique, Burkina Faso, The Gambia.

Available indicators were downloaded from UNAIDs and the WHO Global Health Observatory databases.

In unadjusted analyses, countries with higher prevalences of care seeking for pneumonia and handwashing facilities correlated with lower CCRs ($p<0.01$). Countries with higher prevalence of underweight under-5s correlated with higher CCRs ($p=0.01$). The prevalence of HIV in adults, PCV3 coverage, and the prevalence of biomass fuel use in homes did not correlate with CCRs ($p>0.05$).

After adjusting for the prevalence of care seeking for pneumonia, neither the prevalence of underweight children, nor the prevalence of handwashing remained associated with CCRs. However, analysis included only 125 observations in 38 serotype-specific groups i.e. an average of 3.3. observations per group (range 1-11); so we are unlikely to have had power to determine effects.

Countries with higher prevalence of care seeking for pneumonia have lower CCRs; this may be because this would correlate with more antibiotic use at primary care level and therefore fewer cases presenting to tertiary level hospitals with IPD.

**Supplementary Table 5. The fraction of IPD cases attributable to HIV in the South African population (PAF), by age group[1]; A) using complete cases only (excluding missing data) B) using imputed data where HIV status was missing.**

**A)**

| Age group | RR (95% CI) of association between HIV and IPD | Prevalence of HIV in IPD cases | PAF in % |
|---|---|---|---|
| <5 years | 24.6 (21.0-28.8) | 0.35 (0.33-0.38) | 33.6 (30.6-36.6) |
| 5-14 years | 53.5 (39.1-73.2) | 0.65 (0.61-0.69) | 63.8 (59.7-67.9) |
| 15+ years | 20.7 (18.0-23.9) | 0.80 (0.79-0.82) | 76.1 (74.8-77.5) |

**B)**

| Age group | RR (95% CI) of association between HIV and IPD | Prevalence of HIV in IPD cases | PAF in % |
|---|---|---|---|
| <5 years | 25.6 (21.4-30.7) | 0.34 (0.32-0.37) | 32.7 (32.4-33.8) |
| 5-14 years | 55.0 (43.5-69.5) | 0.62 (0.59-0.66) | 60.9 (59.9-62.9) |
| 15+ years | 20.5 (17.3-24.2) | 0.80 (0.79-0.81) | 76.1 (75.3-77.6) |

[1] We estimated the population attributable fraction (PAF) of HIV on IPD using the formula: $PAF = p_e(RR - 1)/RR$, where 'RR' is the adjusted relative rates of IPD among HIV-infected and uninfected and $p_e$ is the proportion of HIV positive individuals among the IPD cases. The adjusted relative rates were obtained by modelling the count of IPD cases using a negative binomial model.

**Supplementary Table 6. Comparison of calculated CCRs in under-5s in UM/HIC combining pre- and post-PCV data, with previously published CCRs among by Løchen et al. (81)**

| | Gallagher et al. | | Løchen et al. | |
|---|---|---|---|---|
| Serotypes | CCR[1] | 95% CI | CCR | 95% CI |
| PCV7 serotypes | | | | |
| 4 | 143.78 | 72.32, 285.85 | 192.41 | 99.44, 367.35 |
| 6B | 23.58 | 15.89, 34.98 | 75.31 | 45.44, 126.21 |
| 9V | 66.74 | 41.08, 108.43 | 206.87 | 109.36, 391.23 |
| 14 | 62.55 | 39.51, 99.02 | 426.38 | 258.35, 705.69 |
| 18C | 74.44 | 52.28, 106.00 | 273.94 | 147.14, 519.30 |
| 19F | 20.37 | 13.75, 30.19 | 64.66 | 38.85, 107.25 |
| 23F | 19.94 | 14.11, 28.17 | 69.57 | 40.76, 118.35 |
| PCV10 additional types | | | | |
| 1 | 221.85 | 146.43, 336.14 | 1736.98 | 988.83, 3149.16 |
| 5 | 125.22 | 65.07, 240.97 | 732.48 | 295.66, 1858.31 |
| 7F | 812.98 | 286.40, 2307.75 | 958.81 | 564.07, 1672.43 |
| PCV13 additional types | | | | |
| 3 | 48.05 | 30.46, 75.79 | 204.50 | 122.54, 347.60 |
| 6A | 16.56 | 11.50, 23.85 | 43.64 | 25.85, 74.09 |
| 19A | 61.67 | 44.83, 84.85 | 155.42 | 100.70, 241.08 |
| Non-vaccine types | | | | |
| 6C | 15.42 | 6.89, 34.54 | 8.42 | 3.32, 19.71 |
| 7C | 10.99 | 7.86, 15.38 | 39.64 | 11.09, 121.56 |
| 8 | 138.22 | 98.14, 194.66 | 192.82 | 87.85, 413.33 |
| 9N | 23.32 | 15.34, 35.45 | 52.91 | 21.91, 122.30 |
| 10A | 33.14 | 20.44, 53.72 | 72.28 | 40.17, 131.80 |
| 11A | 7.06 | 4.66, 10.71 | 6.76 | 3.12, 13.89 |
| 12F | 94.85 | 57.80, 155.65 | 605.98 | 279.62, 1423.97 |
| 13 | 5.81 | 4.06, 8.32 | 64.50 | 17.87, 208.46 |
| 15A | 17.93 | 11.50, 27.94 | 41.76 | 21.88, 80.11 |
| 16F | 8.57 | 6.33, 11.59 | 39.96 | 18.71, 80.53 |
| 17F | 10.52 | 7.01, 15.78 | 38.61 | 17.47, 82 |
| 19B | 4.91 | 1.36, 17.79 | 1.21 | 0.11, 45.52 |
| 19F | 20.37 | 13.75, 30.19 | 64.66 | 38.85, 107.25 |
| 20 | 7.82 | 4.01, 15.25 | 77.02 | 20.15, 253.36 |
| 21 | 7.74 | 5.34, 11.22 | 20.10 | 8.56, 44.43 |
| 22F | 73.77 | 46.32, 117.49 | 103.46 | 58.24, 187.04 |
| 23A | 7.15 | 4.51, 11.32 | 15.95 | 8.15, 30.53 |
| 23B | 12.71 | 7.02, 23.02 | 26.10 | 13.96, 48.05 |
| 23F | 19.94 | 14.11, 28.17 | 69.57 | 40.76, 118.35 |
| 24F | 52.46 | 35.19, 78.21 | 223.17 | 104.26, 483.25 |
| 28F | 885.93 | 0.14, 5700000 | 61.66 | 5.88, 539.09 |
| 29 | 12.74 | 5.94, 27.32 | 6.42 | 0.30, 49.31 |

| | | | | |
|---|---|---|---|---|
| 31 | 10.76 | 6.42, 18.02 | 49.19 | 16.92, 128.48 |
| 33D | 24.00 | 13.16, 43.76 | | |
| 33F | 90.04 | 54.42, 148.98 | 134.31 | 70.65, 259.75 |
| 34 | 4.67 | 3.05, 7.15 | 10.17 | 2.18, 32.83 |
| 35A | 3.74 | 1.00, 14.05 | 1.49 | 0.12, 57.20 |
| 35B | 13.86 | 10.22, 18.79 | 13.09 | 5.62, 28.07 |
| 35F | 15.16 | 8.06, 28.52 | 15.04 | 6.57, 32.63 |
| 38 | 69.49 | 41.14, 117.37 | 88.26 | 37.78, 199.23 |

[1]The CCRs reported here are from under-5s in UM/HIC but combine pre- and post-PCV data as these are the most comparable CCRs to those published by Løchen et al. which did not disaggregate by PCV period or by country income group but the review included mostly UM/HIC data.

**Supplementary Figure 1. Systematic search terms and search results from searches performed on 14th March 2022 in A) Medline (Ovid SP), B) EMBASE (Ovid SP), and C) the Global Health database**

### A) Medline search

| # ▲ | Searches | Results |
|---|---|---|
| 1 | exp Streptococcus pneumoniae/ or exp Pneumococcal Infections/ or pneumococc*.mp. | 44076 |
| 2 | serotype.mp. or exp Serogroup/ | 40620 |
| 3 | carriage.mp. | 16793 |
| 4 | exp Prevalence/ or prevalen*.mp. | 953946 |
| 5 | 3 or 4 | 965937 |
| 6 | 1 and 2 and 5 | 1564 |
| 7 | exp Pneumonia, Pneumococcal/ or exp Pneumonia, Bacterial/ or exp Pneumonia/ or pneumonia.mp. | 317933 |
| 8 | exp Meningitis, Pneumococcal/ or meningitis.mp. or exp Meningitis/ or exp Meningitis, Bacterial/ | 79775 |
| 9 | invasive pneumococcal disease.mp. | 2735 |
| 10 | bacteremia.mp. or exp Bacteremia/ | 46685 |
| 11 | 7 or 8 or 9 or 10 | 433737 |
| 12 | 1 and 11 | 18706 |
| 13 | surveillance.mp. or Public Health Surveillance/ or Population Surveillance/ or Sentinel Surveillance/ | 262686 |
| 14 | 12 and 13 | 1731 |
| 15 | 6 or 14 | 3065 |

### B) EMBASE search

| # ▲ | Searches | Results |
|---|---|---|
| 1 | Streptococcus pneumoniae.mp. or exp Streptococcus pneumoniae/ | 57012 |
| 2 | pneumococc*.mp. or exp pneumococcal infection/ | 52850 |
| 3 | 1 or 2 | 83757 |
| 4 | serotype.mp. or exp serotype prevalence/ or exp serotype/ | 62757 |
| 5 | carriage.mp. | 21392 |
| 6 | prevalence/ or prevalence.mp. | 1235448 |
| 7 | 5 or 6 | 1251265 |
| 8 | 3 and 4 and 7 | 2009 |
| 9 | exp bacterial pneumonia/ or exp community acquired pneumonia/ or exp pneumonia/ or exp Streptococcus pneumonia/ or pneumonia.mp. | 429602 |
| 10 | exp pneumococcal meningitis/ or meningitis.mp. or exp meningitis/ or exp bacterial meningitis/ | 146842 |
| 11 | invasive pneumococcal disease.mp. | 3557 |
| 12 | exp sepsis/ or bacteremia.mp. or exp bacteremia/ | 324413 |
| 13 | 9 or 10 or 11 or 12 | 828875 |
| 14 | surveillance.mp. or monitoring/ | 510479 |
| 15 | 1 and 13 and 14 | 2065 |
| 16 | 8 or 15 | 3797 |

### C) GLOBAL HEALTH

| # ▲ | Searches | Results |
|---|---|---|
| 1 | Streptococcus pneumoniae.od. or pneumococc*.mp. | 22157 |
| 2 | carriage.mp. | 10736 |
| 3 | serotype.mp. or serotypes/ | 30995 |
| 4 | 1 and 2 and 3 | 792 |
| 5 | exp bacterial pneumonia/ or exp pneumonia/ or pneumonia.mp. or exp community acquired pneumonia/ | 56204 |
| 6 | exp bacterial meningitis/ or meningitis.mp. or exp meningitis/ | 26228 |
| 7 | invasive pneumococcal disease.mp. | 1971 |
| 8 | bacteremia.mp. or bacteraemia.sh. | 16600 |
| 9 | 5 or 6 or 7 or 8 | 94450 |
| 10 | exp surveillance/ or surveillance.mp. or exp sentinel surveillance/ or exp syndromic surveillance/ | 151836 |
| 11 | 1 and 9 and 10 | 1529 |
| 12 | 4 or 11 | 2240 |

**Supplementary Figure 2. Derivation of the Confidence Intervals for Case-Carrier Ratios**

Let $\hat{\lambda}$ denote the estimated IPD incidence, which is calculated as

$$\hat{\lambda} = d/y\ ,$$

where $d$ is the number of IPD cases and $y$ is the number of person years of surveillance.

Let $\hat{p}$ denote the estimated carriage prevalence, which is calculated

$$\hat{p} = x/n,$$

where $x$ is the number of carriers and $n$ is the number of participants in the carriage survey.

Then the estimated case carrier ratio is given by:

$$\widehat{CCR} = \hat{\lambda}/\hat{p}.$$

To obtain a confidence interval for $\widehat{CCR}$, we first compute a confidence interval for $\log\widehat{CCR}$ and then the exponentiate the lower and upper bounds of this interval.

The formula for $\mathrm{var}[\log\widehat{CCR}]$ is derived via the delta method as follows:

$$\mathrm{var}\left[\log\widehat{CCR}\right] = \frac{\mathrm{var}(\hat{\lambda})}{\hat{\lambda}^2} + \frac{\mathrm{var}(\hat{p})}{\hat{p}^2}$$

$$= \frac{d/y^2}{\left(\frac{d}{y}\right)^2} + \frac{\hat{p}(1-\hat{p})/n}{\hat{p}^2}$$

$$= \frac{1}{d} + \frac{(1-\hat{p})}{n\hat{p}}\ .$$

Therefore, the standard error for $\log\widehat{CCR}$ is given by:

$$\mathrm{SE}(\log\widehat{CCR}) = \sqrt{\frac{1}{d} + \frac{(1-\hat{p})}{n\hat{p}}}.$$

The 95% confidence interval for $\log\widehat{CCR}$ is calculated in the usual way as:

$$\log\widehat{CCR} \pm 1.96 \times \ \mathrm{SE}(\log\widehat{CCR})$$

**Supplementary Figure 3. Comparison of CCRs pre-post PCV introduction, by age and by country income group**

**A) CCRs post- vs. pre-PCV introduction, in under-5s in LMIC**

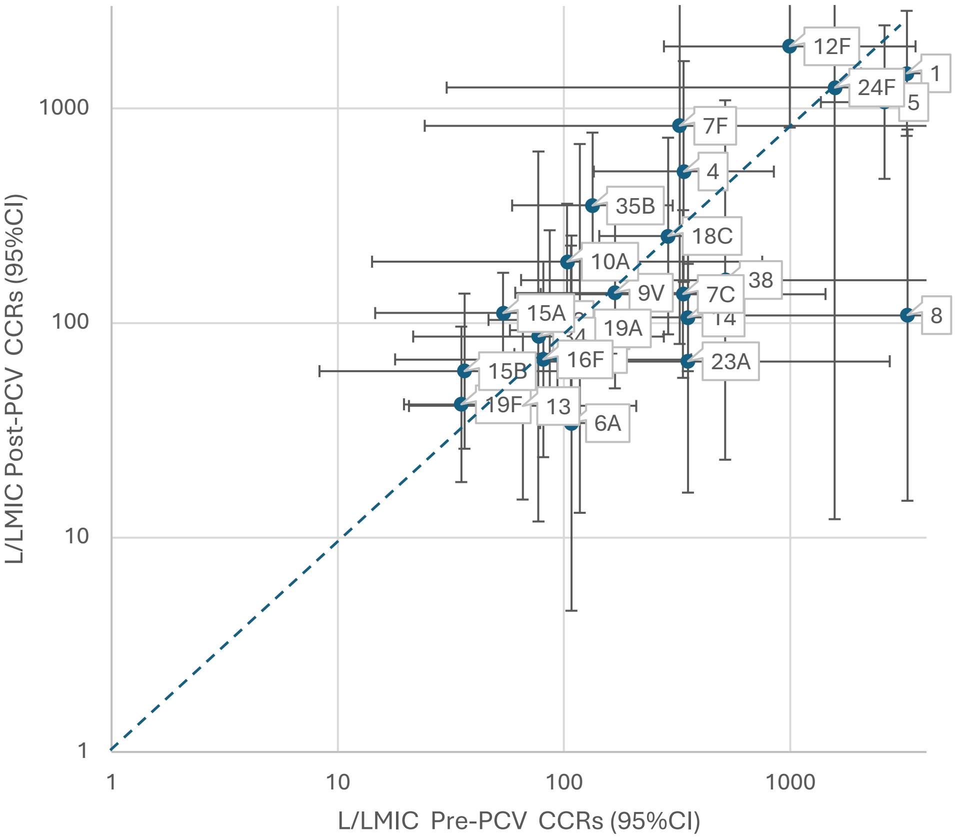

**B) CCRs pre- vs. post PCV introduction, in under-5s in UM/HIC**

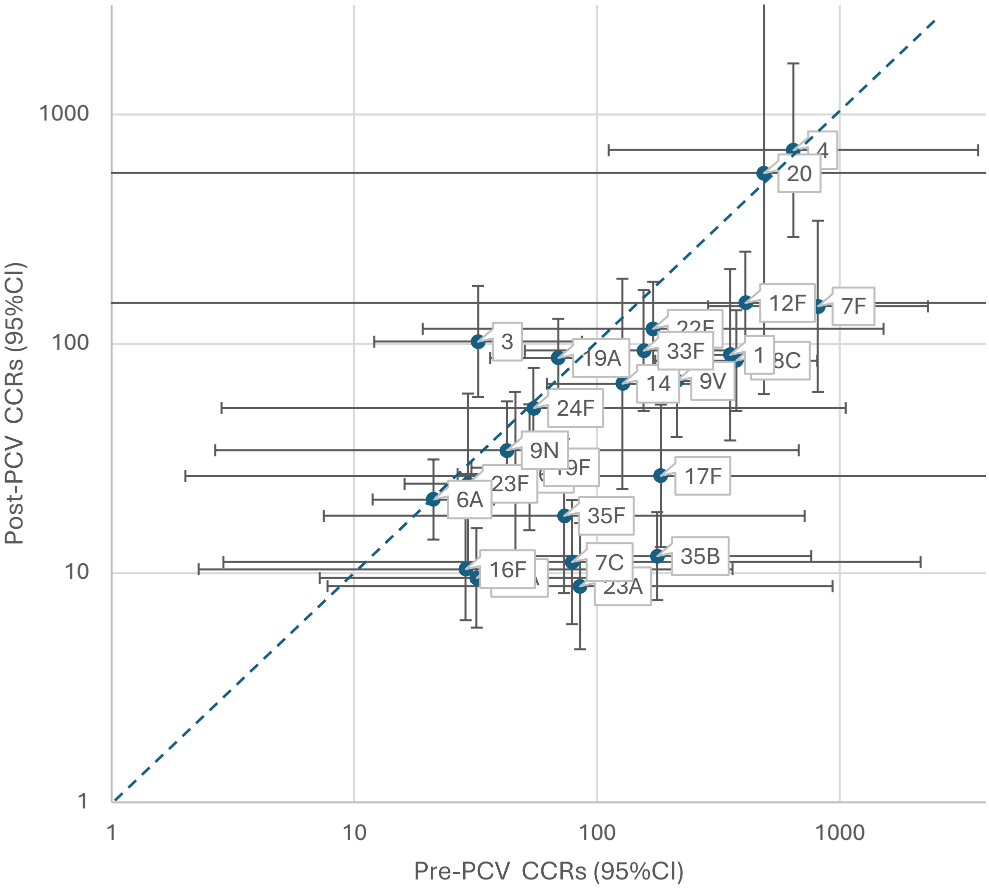

**C) CCRs pre- vs. post PCV introduction, in 5-14 year olds in LMIC**

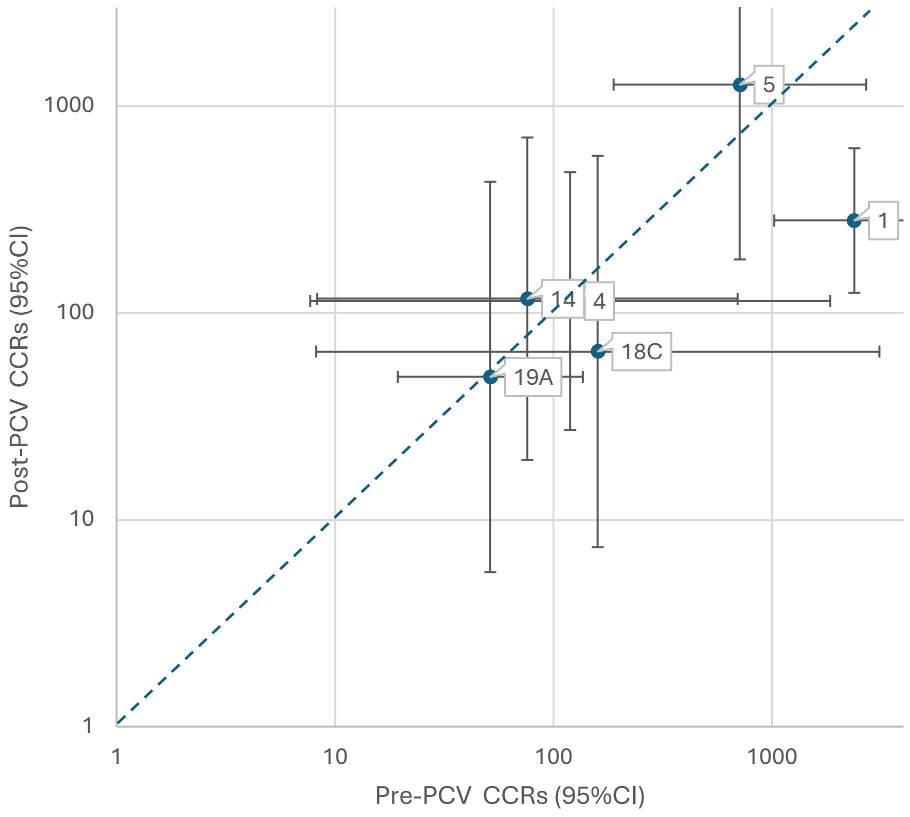

**D) CCRs pre- vs. post PCV introduction, in 5-14 year olds in UM/HIC**

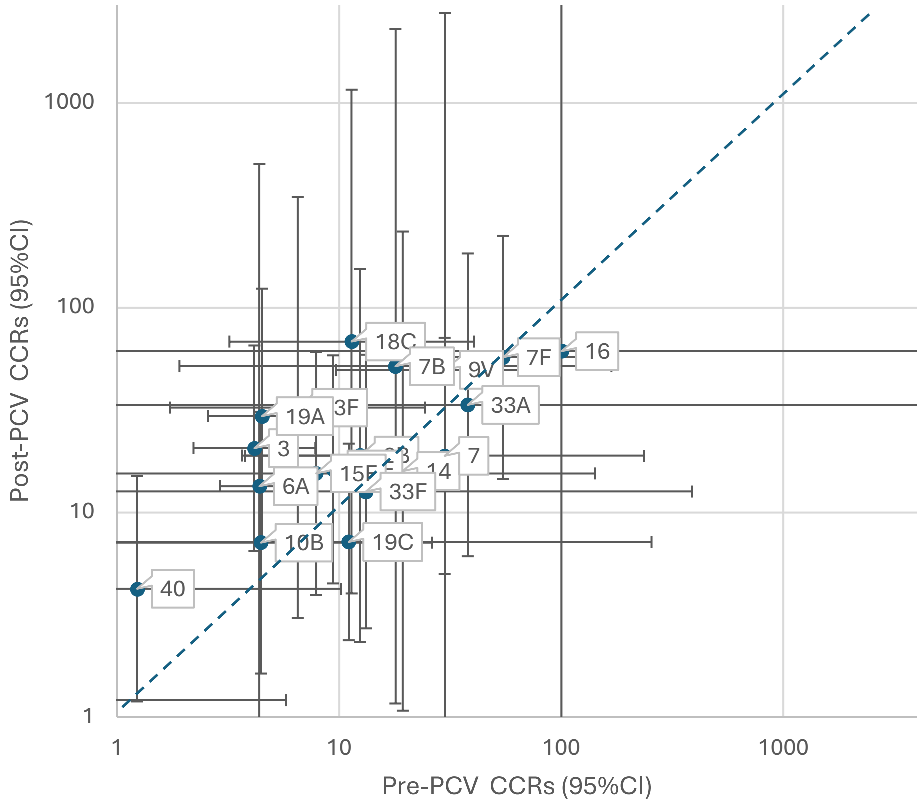

**E) CCRs pre- vs. post PCV introduction, in 15 years and above in LMIC**

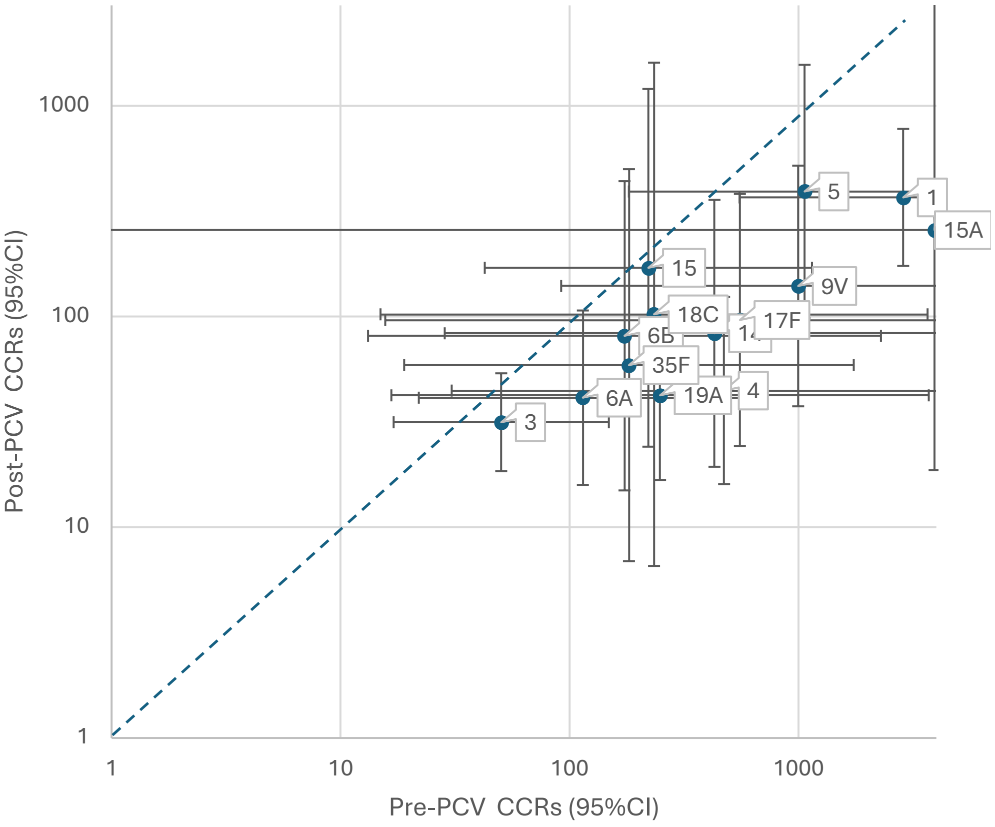

**F) CCRs pre- vs. post PCV introduction, in 15 year olds and above in UM/HIC**

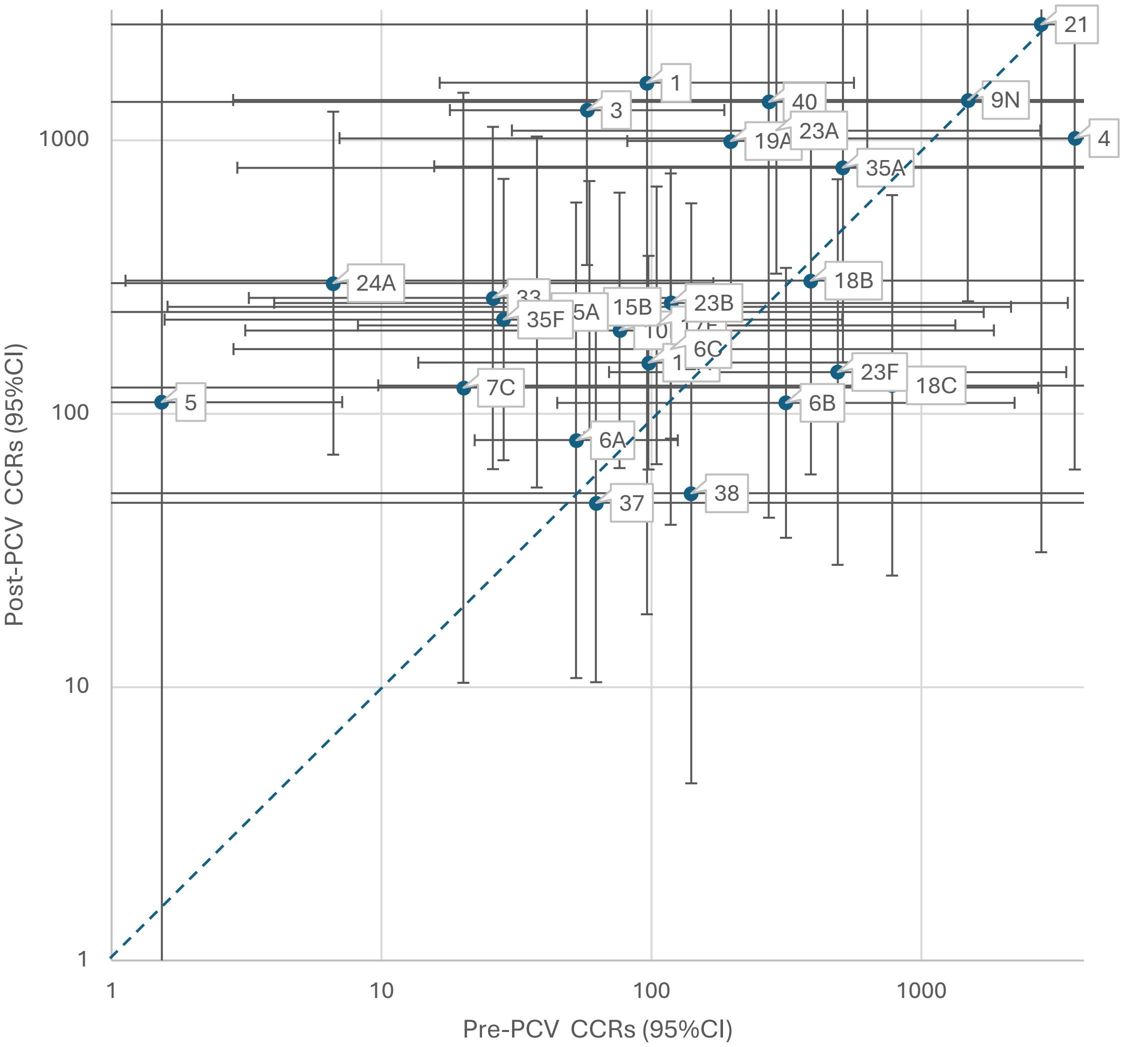

**Supplementary Figure 4. Analysis of CCRs among children under-5 years of age, over time (restricted to datasets containing calendar year relative to PCV introduction)**

**A) In LMICs, PCV7 VTs**

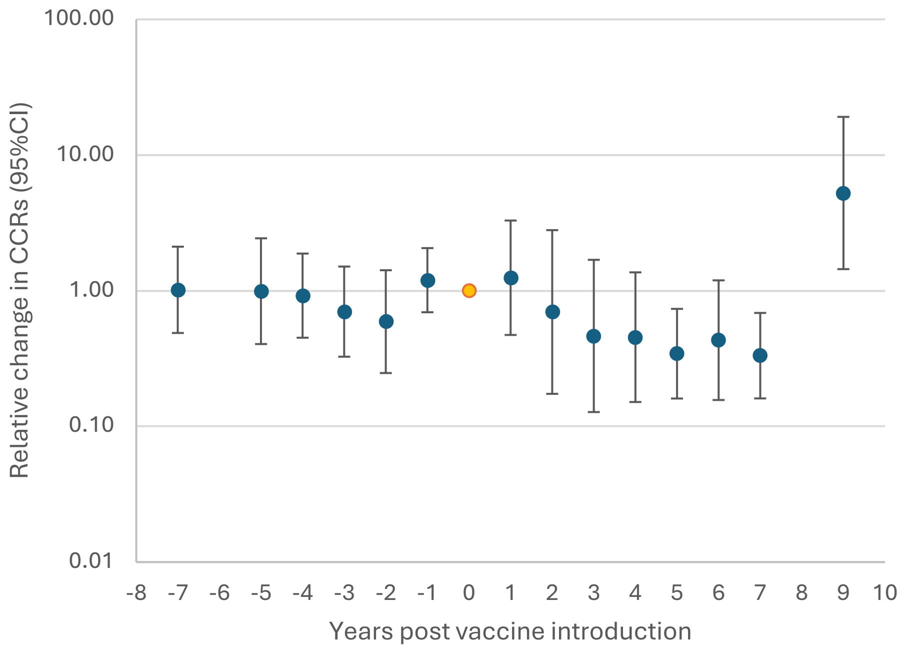

**B) In LMICs, NVTs**

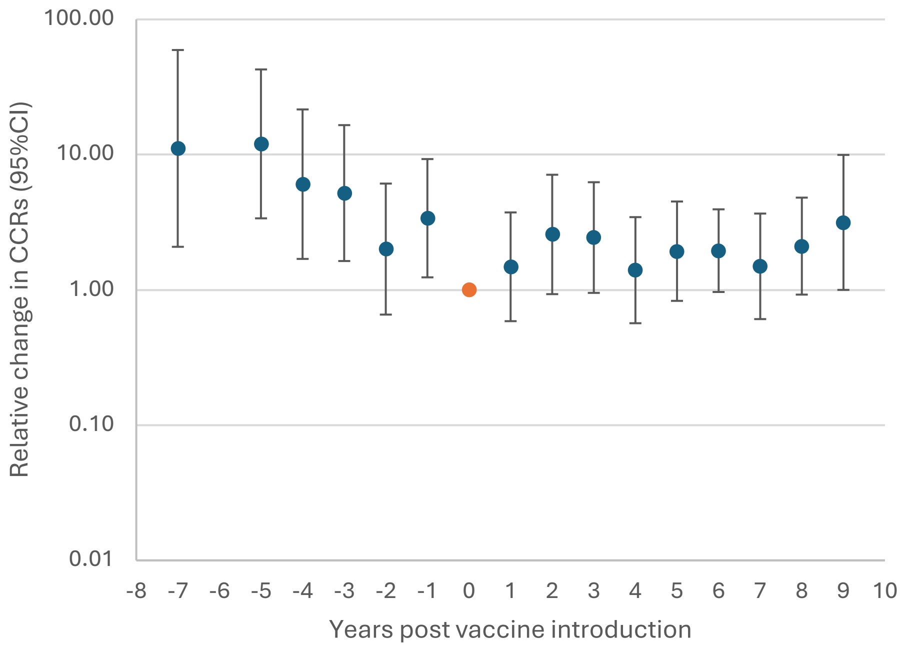

**C) In UM/HICs (excluding South Africa), PCV7 VTs**

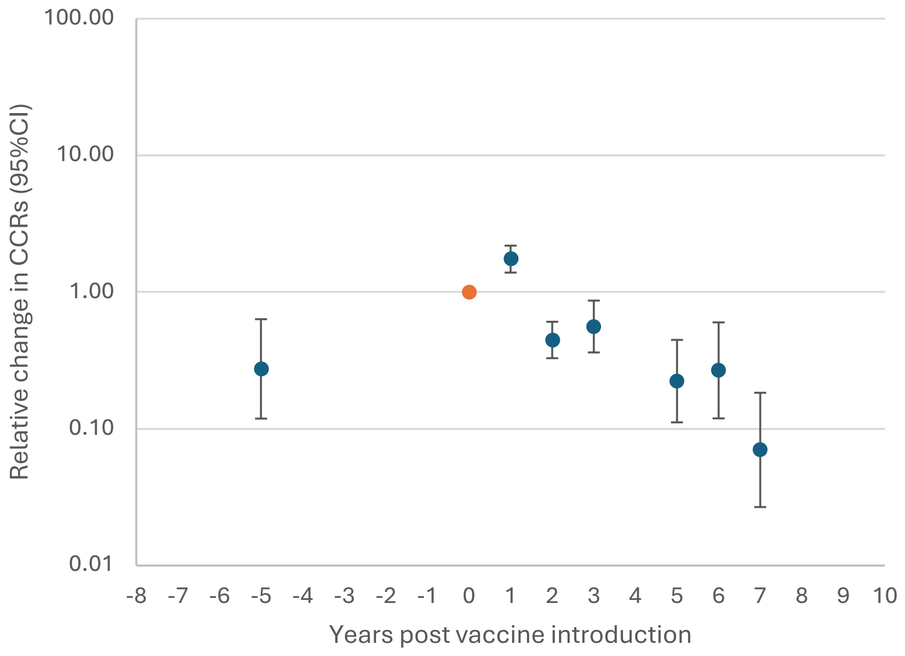

**D) In UM/HICs (excluding South Africa), NVTs**

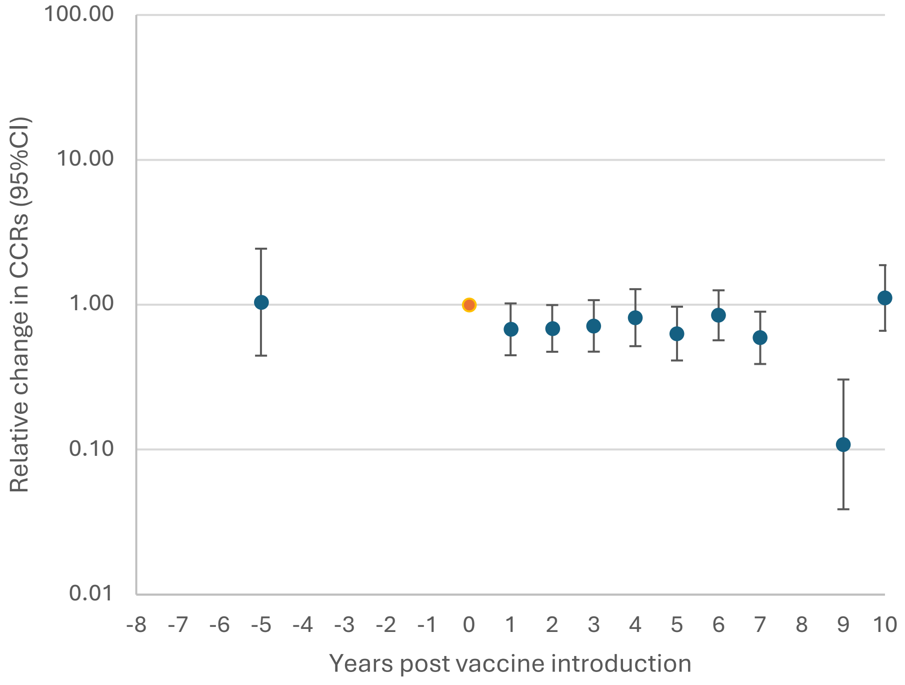

E) **In South African data (combining HIV infected and HIV uninfected populations), PCV7 VTs**

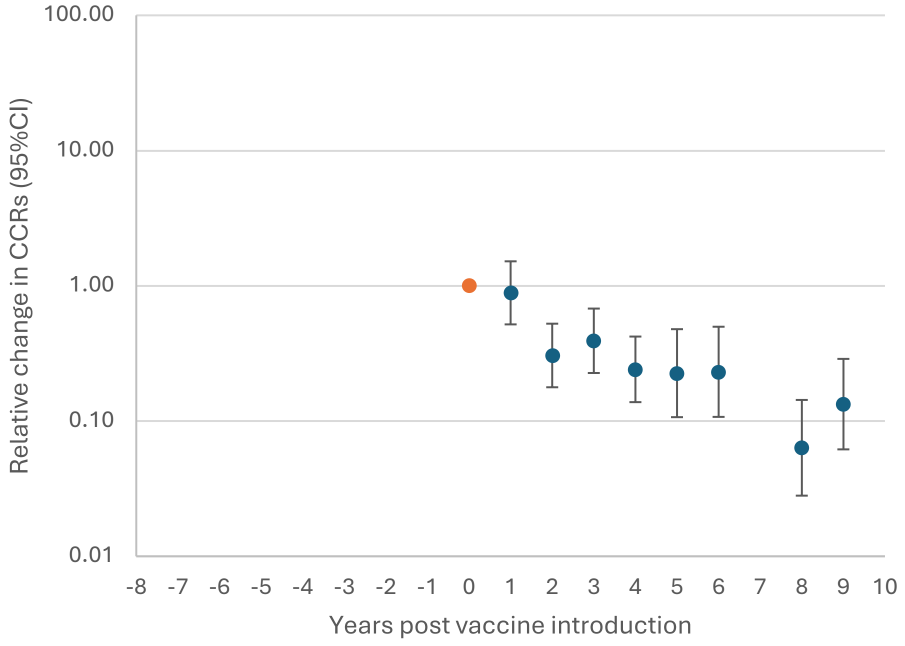

F) **In South African data (combining HIV infected and HIV uninfected populations), NVTs**

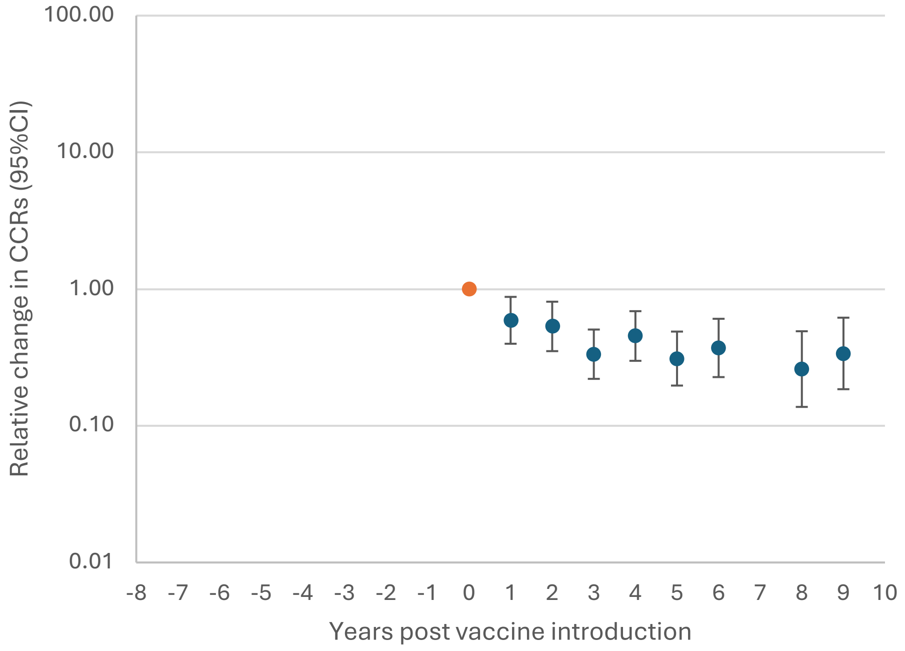

**Supplementary Figure 5. CCRs by sex in A) children under 5 years of age, B) children between 5 and 14 years of age, C) older children and adults 15 years of age and over.**

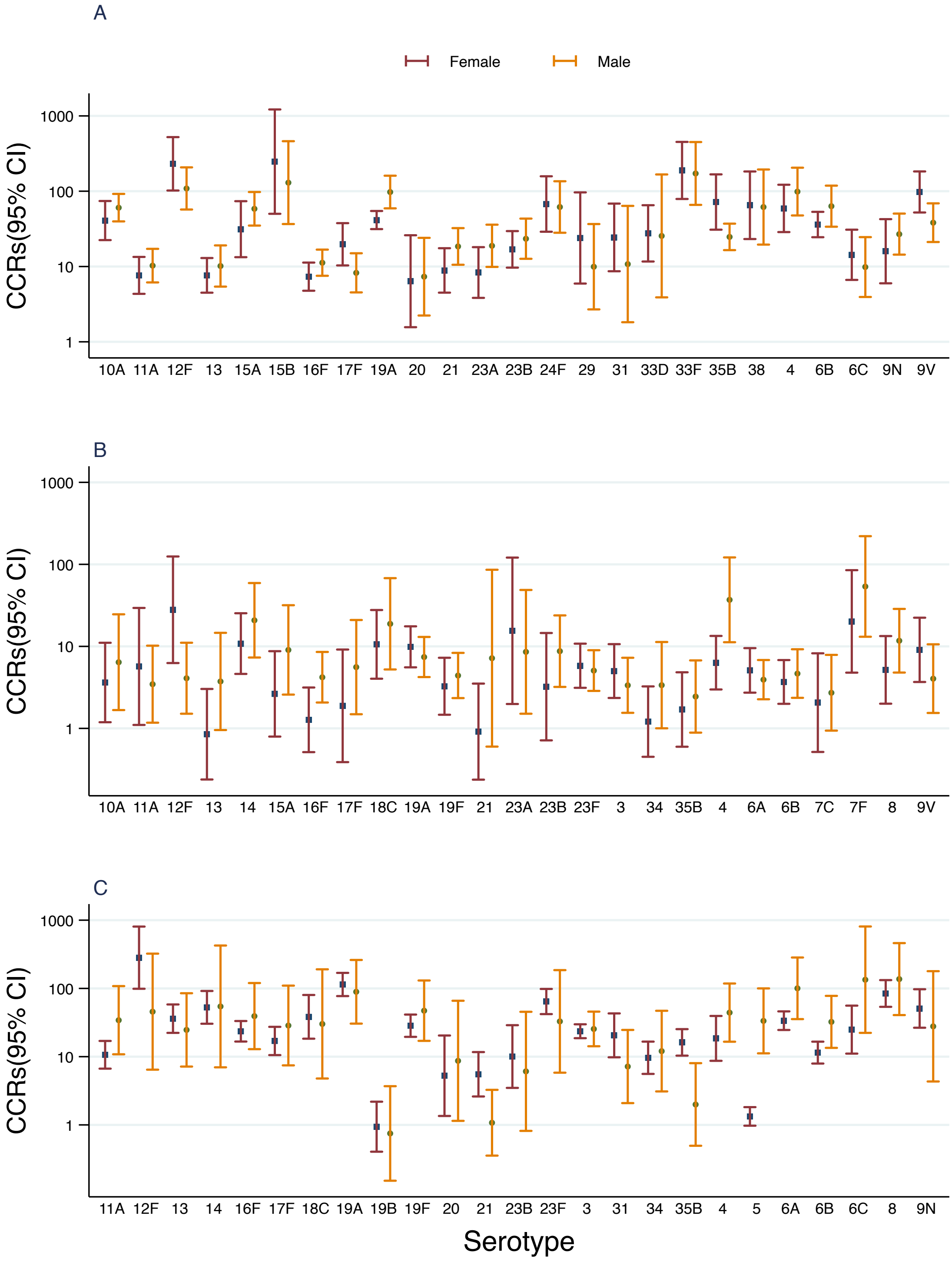

**Supplementary Figure 6. CCRs by HIV status using imputed data where HIV status of IPD cases was missing[1]; A) children under 5 years of age, B) children between 5 and 14 years of age, C) older children and adults 15 years of age and over**

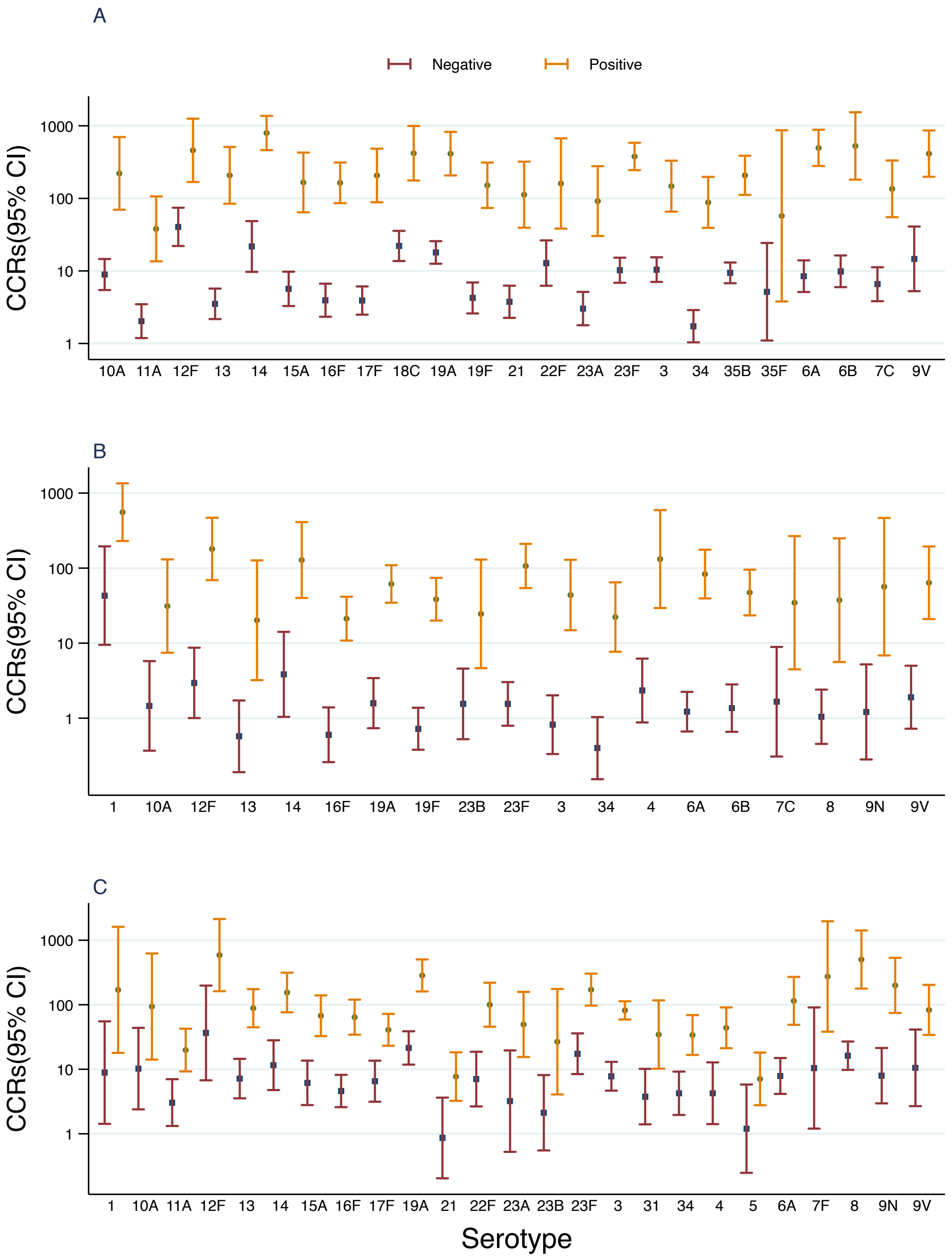

[1] The percentage of IPD cases missing HIV status was high (55% in children under-5 years of age, 63% in 5-14 year olds, and 68% in 15 year olds and above)

### References

1. Desmet S, Lagrou K, Wyndham-Thomas C, Braeye T, Verhaegen J, Maes P, et al. Dynamic changes in paediatric invasive pneumococcal disease after sequential switches of conjugate vaccine in Belgium: a national retrospective observational study. The Lancet Infectious diseases. 2021;21(1):127-36.
2. Wouters I, Desmet S, Van Heirstraeten L, Blaizot S, Verhaegen J, Van Damme P, et al. Follow-up of serotype distribution and antimicrobial susceptibility of Streptococcus pneumoniae in child carriage after a PCV13-to-PCV10 vaccine switch in Belgium. Vaccine. 2019;37(8):1080-6.
3. Desmet S, Wouters I, Heirstraeten LV, Beutels P, Van Damme P, Malhotra-Kumar S, et al. In-depth analysis of pneumococcal serotypes in Belgian children (2015-2018): Diversity, invasive disease potential, and antimicrobial susceptibility in carriage and disease. Vaccine. 2021;39(2):372-9.
4. Harboe ZB, Slotved HC, Konradsen HB, Kaltoft MS. A Pneumococcal Carriage Study in Danish Pre-school Children before the Introduction of Pneumococcal Conjugate Vaccination. Open Microbiol J. 2012;6:40-4.
5. Harboe ZB, Valentiner-Branth P, Benfield TL, Christensen JJ, Andersen PH, Howitz M, et al. Early effectiveness of heptavalent conjugate pneumococcal vaccination on invasive pneumococcal disease after the introduction in the Danish Childhood Immunization Programme. Vaccine. 2010;28(14):2642-7.
6. Hussain M, Melegaro A, Pebody RG, George R, Edmunds WJ, Talukdar R, et al. A longitudinal household study of Streptococcus pneumoniae nasopharyngeal carriage in a UK setting. Epidemiol Infect. 2005;133(5):891-8.
7. Flasche S, Hoek AJV, Sheasby E, Waight P, Andrews N, George R. Effect of pneumococcal conjugate vaccination on serotype-specific carriage and invasive disease in England: a cross-sectional study. PLoS medicine. 2011;8.
8. Kent A, Makwana A, Sheppard CL, Collins S, Fry NK, Heath PT, et al. Invasive Pneumococcal Disease in UK Children <1 Year of Age in the Post-13-Valent Pneumococcal Conjugate Vaccine Era: What Are the Risks Now? Clinical infectious diseases : an official publication of the Infectious Diseases Society of America. 2019;69(1):84-90.
9. Foster D, Knox K, Walker AS, Griffiths DT, Moore H, Haworth E, et al. Invasive pneumococcal disease: epidemiology in children and adults prior to implementation of the conjugate vaccine in the Oxfordshire region, England. J Med Microbiol. 2008;57(Pt 4):480-7.
10. Johnson AP, Waight P, Andrews N, Pebody R, George RC, Miller E. Morbidity and mortality of pneumococcal meningitis and serotypes of causative strains prior to introduction of the 7-valent conjugant pneumococcal vaccine in England. J Infect. 2007;55(5):394-9.
11. van Hoek AJ, Sheppard CL, Andrews NJ, Waight PA, Slack MP, Harrison TG, et al. Pneumococcal carriage in children and adults two years after introduction of the thirteen valent pneumococcal conjugate vaccine in England. Vaccine. 2014;32(34):4349-55.
12. Makwana A, Sheppard C, Borrow R, Fry N, Andrews NJ, Ladhani SN. Characteristics of Children With Invasive Pneumococcal Disease After the Introduction of the 13-valent Pneumococcal Conjugate Vaccine in England and Wales, 2010-2016. The Pediatric infectious disease journal. 2018;37(7):697-703.
13. Southern J, Andrews N, Sandu P, Sheppard CL, Waight PA, Fry NK, et al. Pneumococcal carriage in children and their household contacts six years after introduction of the 13-valent pneumococcal conjugate vaccine in England. PloS one. 2018;13(5):e0195799.
14. Ladhani SN, Collins S, Djennad A, Sheppard CL, Borrow R, Fry NK, et al. Rapid increase in non-vaccine serotypes causing invasive pneumococcal disease in England and Wales, 2000–17: a prospective national observational cohort study. The Lancet Infectious Diseases. 2018;18(4):441-51.
15. Kandasamy R, Voysey M, Collins S, Berbers G, Robinson H, Noel I, et al. Persistent Circulation of Vaccine Serotypes and Serotype Replacement After 5 Years of Infant Immunization With 13-Valent Pneumococcal Conjugate Vaccine in the United Kingdom. The Journal of infectious diseases. 2020;221(8):1361-70.
16. Dunais B, Bruno-Bazureault P, Carsenti-Dellamonica H, Touboul P, Pradier C. A decade-long surveillance of nasopharyngeal colonisation with Streptococcus pneumoniae among children attending day-care centres in south-eastern France: 1999-2008. Eur J Clin Microbiol Infect Dis. 2011;30(7):837-43.
17. Lepoutre A, Varon E, Georges S, Gutmann L, Lévy-Bruhl D. Impact of infant pneumococcal vaccination on invasive pneumococcal diseases in France, 2001-2006. Euro surveillance : bulletin Europeen sur les maladies transmissibles = European communicable disease bulletin. 2008;13(35).
18. Dunais B, Bruno P, Touboul P, Degand N, Sakarovitch C, Fontas E, et al. Impact of the 13-valent pneumococcal conjugate vaccine on nasopharyngeal carriage of Streptococcus pneumoniae among children attending group daycare in southeastern France. The Pediatric infectious disease journal. 2015;34(3):286-8.
19. Lepoutre A, Varon E, Georges S, Dorléans F, Janoir C, Gutmann L, et al. Impact of the pneumococcal conjugate vaccines on invasive pneumococcal disease in France, 2001-2012. Vaccine. 2015;33(2):359-66.
20. Nikolova KA, Andersson M, Slotved HC, Koch A. Effectiveness of the 13-Valent Pneumococcal Conjugate Vaccine on Invasive Pneumococcal Disease in Greenland. Vaccines (Basel). 2021;9(10).

21. Navne JE, Koch A, Slotved HC, Andersson M, Melbye M, Ladefoged K, et al. Effect of the 13-valent pneumococcal conjugate vaccine on nasopharyngeal carriage by respiratory pathogens among Greenlandic children. Int J Circumpolar Health. 2017;76(1):1309504.
22. Hjálmarsdóttir M, Quirk SJ, Haraldsson G, Erlendsdóttir H, Haraldsson Á, Kristinsson KG. Comparison of Serotype Prevalence of Pneumococci Isolated from Middle Ear, Lower Respiratory Tract and Invasive Disease Prior to Vaccination in Iceland. PloS one. 2017;12(1):e0169210.
23. Quirk SJ, Haraldsson G, Erlendsdóttir H, Hjálmarsdóttir M, van Tonder AJ, Hrafnkelsson B, et al. Effect of Vaccination on Pneumococci Isolated from the Nasopharynx of Healthy Children and the Middle Ear of Children with Otitis Media in Iceland. Journal of clinical microbiology. 2018;56(12).
24. Vestrheim DF, Høiby EA, Aaberge IS, Caugant DA. Impact of a pneumococcal conjugate vaccination program on carriage among children in Norway. Clinical and vaccine immunology : CVI. 2010;17(3):325-34.
25. Vestrheim DF, Løvoll O, Aaberge IS, Caugant DA, Høiby EA, Bakke H, et al. Effectiveness of a 2+1 dose schedule pneumococcal conjugate vaccination programme on invasive pneumococcal disease among children in Norway. Vaccine. 2008;26(26):3277-81.
26. Steens A, Bergsaker MA, Aaberge IS, Rønning K, Vestrheim DF. Prompt effect of replacing the 7-valent pneumococcal conjugate vaccine with the 13-valent vaccine on the epidemiology of invasive pneumococcal disease in Norway. Vaccine. 2013;31(52):6232-8.
27. Rodrigues F, Foster D, Caramelo F, Serranho P, Gonçalves G, Januário L, et al. Progressive changes in pneumococcal carriage in children attending daycare in Portugal after 6 years of gradual conjugate vaccine introduction show falls in most residual vaccine serotypes but no net replacement or trends in diversity. Vaccine. 2012;30(26):3951-6.
28. Nunes S, Félix S, Valente C, Simões AS, Tavares DA, Almeida ST, et al. The impact of private use of PCV7 in 2009 and 2010 on serotypes and antimicrobial resistance of Streptococcus pneumoniae carried by young children in Portugal: Comparison with data obtained since 1996 generating a 15-year study prior to PCV13 introduction. Vaccine. 2016;34(14):1648-56.
29. Aguiar SI, Brito MJ, Horacio AN, Lopes JP, Ramirez M, Melo-Cristino J. Decreasing incidence and changes in serotype distribution of invasive pneumococcal disease in persons aged under 18 years since introduction of 10-valent and 13-valent conjugate vaccines in Portugal, July 2008 to June 2012. Euro surveillance : bulletin Europeen sur les maladies transmissibles = European communicable disease bulletin. 2014;19(12):20750.
30. Félix S, Handem S, Nunes S, Paulo AC, Candeias C, Valente C, et al. Impact of private use of the 13-valent pneumococcal conjugate vaccine (PCV13) on pneumococcal carriage among Portuguese children living in urban and rural regions. Vaccine. 2021;39(32):4524-33.
31. Silva-Costa C, Brito MJ, Aguiar SI, Lopes JP, Ramirez M, Melo-Cristino J. Dominance of vaccine serotypes in pediatric invasive pneumococcal infections in Portugal (2012-2015). Scientific reports. 2019;9(1):6.
32. Silva-Costa C, Gomes-Silva J, Prados L, Ramirez M, Melo-Cristino J, On Behalf Of The Portuguese Group For The Study Of Streptococcal I, et al. Pediatric Invasive Pneumococcal Disease Three Years after PCV13 Introduction in the National Immunization Plan-The Continued Importance of Serotype 3. Microorganisms. 2021;9(7).
33. Desai AP, Sharma D, Crispell EK, Baughman W, Thomas S, Tunali A, et al. Decline in Pneumococcal Nasopharyngeal Carriage of Vaccine Serotypes After the Introduction of the 13-Valent Pneumococcal Conjugate Vaccine in Children in Atlanta, Georgia. The Pediatric infectious disease journal. 2015;34(11):1168-74.
34. Sharma D, Baughman W, Holst A, Thomas S, Jackson D, Gloria CM. Pneumococcal carriage and invasive disease in children before introduction of the 13-valent conjugate vaccine: comparison with the era before 7-valent conjugate vaccine. The Pediatric infectious disease journal. 2013;32.
35. Beall B, Chochua S, Gertz RE, Jr., Li Y, Li Z, McGee L, et al. A Population-Based Descriptive Atlas of Invasive Pneumococcal Strains Recovered Within the U.S. During 2015-2016. Front Microbiol. 2018;9:2670.
36. Schroeder MR, Chancey ST, Thomas S, Kuo WH, Satola SW, Farley MM, et al. A Population-Based Assessment of the Impact of 7- and 13-Valent Pneumococcal Conjugate Vaccines on Macrolide-Resistant Invasive Pneumococcal Disease: Emergence and Decline of Streptococcus pneumoniae Serotype 19A (CC320) With Dual Macrolide Resistance Mechanisms. Clinical infectious diseases : an official publication of the Infectious Diseases Society of America. 2017;65(6):990-8.
37. Moore MR, Link-Gelles R, Schaffner W, Lynfield R, Lexau C, Bennett NM, et al. Effect of use of 13-valent pneumococcal conjugate vaccine in children on invasive pneumococcal disease in children and adults in the USA: analysis of multisite, population-based surveillance. The Lancet Infectious diseases. 2015;15(3):301-9.
38. Gierke R, Farley MM, Schaffner W, Thomas A, Reingold A, Harrison L, et al. 1299. Epidemiology of Invasive Pneumococcal Disease (IPD) in the United States 2011-2019. Open Forum Infect Dis. 2021;8(Supplement_1):S737-S8.
39. Motlova J, Benes C, Kriz P. Incidence of invasive pneumococcal disease in the Czech Republic and serotype coverage by vaccines, 1997-2006. Epidemiol Infect. 2009;137(4):562-9.

40. Zemlicková H, Urbásková P, Adámková V, Motlová J, Lebedová V, Procházka B. Characteristics of Streptococcus pneumoniae, Haemophilus influenzae, Moraxella catarrhalis and Staphylococcus aureus isolated from the nasopharynx of healthy children attending day-care centres in the Czech Republic. Epidemiol Infect. 2006;134(6):1179-87.
41. Andrade AL, Ternes YM, Vieira MA, Moreira WG, Lamaro-Cardoso J, Kipnis A, et al. Direct effect of 10-valent conjugate pneumococcal vaccination on pneumococcal carriage in children Brazil. PloS one. 2014;9(6):e98128.
42. Agudelo CI, Castañeda-Orjuela C, Brandileone MCC, Echániz-Aviles G, Almeida SCG, Carnalla-Barajas MN, et al. The direct effect of pneumococcal conjugate vaccines on invasive pneumococcal disease in children in the Latin American and Caribbean region (SIREVA 2006-17): a multicentre, retrospective observational study. The Lancet Infectious diseases. 2021;21(3):405-17.
43. Toledo ME, Casanova MF, Linares-Pérez N, García-Rivera D, Toraño Peraza G, Barcos Pina I, et al. Prevalence of Pneumococcal Nasopharyngeal Carriage Among Children 2-18 Months of Age: Baseline Study Pre Introduction of Pneumococcal Vaccination in Cuba. The Pediatric infectious disease journal. 2017;36(1):e22-e8.
44. Nunes MC, Jones SA, Groome MJ, Kuwanda L, Van Niekerk N, von Gottberg A, et al. Acquisition of Streptococcus pneumoniae in South African children vaccinated with 7-valent pneumococcal conjugate vaccine at 6, 14 and 40 weeks of age. Vaccine. 2015;33(5):628-34.
45. Tempia S, Wolter N, Cohen C, Walaza S, von Mollendorf C, Cohen AL, et al. Assessing the impact of pneumococcal conjugate vaccines on invasive pneumococcal disease using polymerase chain reaction-based surveillance: an experience from South Africa. BMC infectious diseases. 2015;15:450.
46. von Gottberg A, de Gouveia L, Tempia S, Quan V, Meiring S, von Mollendorf C, et al. Effects of Vaccination on Invasive Pneumococcal Disease in South Africa. N Engl J Med. 2014;371(20):1889-99.
47. von Mollendorf C, Cohen C, Tempia S, Meiring S, de Gouveia L, Quan V, et al. Epidemiology of Serotype 1 Invasive Pneumococcal Disease, South Africa, 2003-2013. Emerging infectious diseases. 2016;22(2):261-70.
48. Ndlangisa K, du Plessis M, Allam M, Wolter N, de Gouveia L, Klugman KP, et al. Invasive Disease Caused Simultaneously by Dual Serotypes of Streptococcus pneumoniae. Journal of clinical microbiology. 2018;56(1).
49. Skosana Z, Von Gottberg A, Olorunju S, Mohale T, Du Plessis M, Adams T, et al. Non-vaccine serotype pneumococcal carriage in healthy infants in South Africa following introduction of the 13-valent pneumococcal conjugate vaccine. S Afr Med J. 2021;111(2):143-8.
50. Madhi SA, Nzenze SA, Nunes MC, Chinyanganya L, Van Niekerk N, Kahn K, et al. Residual colonization by vaccine serotypes in rural South Africa four years following initiation of pneumococcal conjugate vaccine immunization. Expert Rev Vaccines. 2020;19(4):383-93.
51. Nzenze SA, Shiri T, Nunes MC, Klugman KP, Kahn K, Twine R, et al. Temporal changes in pneumococcal colonization in a rural African community with high HIV prevalence following routine infant pneumococcal immunization. The Pediatric infectious disease journal. 2013;32(11):1270-8.
52. Hammitt LL, Etyang AO, Morpeth SC, Ojal J, Mutuku A, Mturi N, et al. Effect of ten-valent pneumococcal conjugate vaccine on invasive pneumococcal disease and nasopharyngeal carriage in Kenya: a longitudinal surveillance study. Lancet (London, England). 2019;393(10186):2146-54.
53. Hammitt LL, Akech DO, Morpeth SC, Karani A, Kihuha N, Nyongesa S. Population effect of 10-valent pneumococcal conjugate vaccine on nasopharyngeal carriage of *Streptococcus pneumoniae* and non-typeable *Haemophilus influenzae* in Kilifi, Kenya: findings from cross-sectional carriage studies. Lancet Glob Heal. 2014;2.
54. Abdullahi O, Karani A, Tigoi CC, Mugo D, Kungu S, Wanjiru E, et al. The prevalence and risk factors for pneumococcal colonization of the nasopharynx among children in Kilifi District, Kenya. PloS one. 2012;7(2):e30787.
55. Soeters HM, Kambiré D, Sawadogo G, Ouédraogo-Traoré R, Bicaba B, Medah I, et al. Impact of 13-Valent Pneumococcal Conjugate Vaccine on Pneumococcal Meningitis, Burkina Faso, 2016-2017. The Journal of infectious diseases. 2019;220(220 Suppl 4):S253-s62.
56. Kaboré L, Adebanjo T, Njanpop-Lafourcade BM, Ouangraoua S, Tarbangdo FT, Meda B, et al. Pneumococcal Carriage in Burkina Faso After 13-Valent Pneumococcal Conjugate Vaccine Introduction: Results From 2 Cross-sectional Population-Based Surveys. The Journal of infectious diseases. 2021;224(12 Suppl 2):S258-s66.
57. Chaguza C, Senghore M, Bojang E, Lo SW, Ebruke C, Gladstone RA, et al. Carriage Dynamics of Pneumococcal Serotypes in Naturally Colonized Infants in a Rural African Setting During the First Year of Life. Front Pediatr. 2020;8:587730.
58. Odutola A, Antonio M, Owolabi O, Bojang A, Foster-Nyarko E, Donkor S, et al. Comparison of the prevalence of common bacterial pathogens in the oropharynx and nasopharynx of gambian infants. PloS one. 2013;8(9):e75558.
59. Sanneh B, Okoi C, Grey-Johnson M, Bah-Camara H, Kunta Fofana B, Baldeh I, et al. Declining Trends of Pneumococcal Meningitis in Gambian Children After the Introduction of Pneumococcal Conjugate Vaccines.

Clinical infectious diseases : an official publication of the Infectious Diseases Society of America. 2019;69(Suppl 2):S126-s32.
60. Mackenzie GA, Hill PC, Jeffries DJ, Hossain I, Uchendu U, Ameh D, et al. Effect of the introduction of pneumococcal conjugate vaccination on invasive pneumococcal disease in The Gambia: a population-based surveillance study. The Lancet Infectious Diseases. 2016;16(6):703-11.
61. Roca A, Bojang A, Bottomley C, Gladstone RA, Adetifa JU, Egere U, et al. Effect on nasopharyngeal pneumococcal carriage of replacing PCV7 with PCV13 in the Expanded Programme of Immunization in The Gambia. Vaccine. 2015;33(51):7144-51.
62. Usen S, Adegbola R, Mulholland K, Jaffar S, Hilton S, Oparaugo A, et al. Epidemiology of invasive pneumococcal disease in the Western Region, The Gambia. The Pediatric infectious disease journal. 1998;17(1):23-8.
63. Mackenzie GA, Hill PC, Jeffries DJ, Ndiaye M, Sahito SM, Hossain I, et al. Impact of the introduction of pneumococcal conjugate vaccination on invasive pneumococcal disease and pneumonia in The Gambia: 10 years of population-based surveillance. The Lancet Infectious diseases. 2021;21(9):1293-302.
64. Hill PC, Akisanya A, Sankareh K, Cheung YB, Saaka M, Lahai G, et al. Nasopharyngeal carriage of Streptococcus pneumoniae in Gambian villagers. Clinical infectious diseases : an official publication of the Infectious Diseases Society of America. 2006;43(6):673-9.
65. Hill PC, Townend J, Antonio M, Akisanya B, Ebruke C, Lahai G, et al. Transmission of Streptococcus pneumoniae in rural Gambian villages: a longitudinal study. Clinical infectious diseases : an official publication of the Infectious Diseases Society of America. 2010;50(11):1468-76.
66. Usuf E, Badji H, Bojang A, Jarju S, Ikumapayi UN, Antonio M, et al. Pneumococcal carriage in rural Gambia prior to the introduction of pneumococcal conjugate vaccine: a population-based survey. Trop Med Int Health. 2015;20(7):871-9.
67. Usuf E, Bottomley C, Gladstone R, Bojang E, Jawneh K, Cox I, et al. Persistent and Emerging Pneumococcal Carriage Serotypes in a Rural Gambian Community After 10 Years of Pneumococcal Conjugate Vaccine Pressure. Clinical infectious diseases : an official publication of the Infectious Diseases Society of America. 2021;73(11):e3825-e35.
68. Ashu EE, Jarju S, Dione M, Mackenzie G, Ikumapayi UN, Manjang A, et al. Population structure, epidemiology and antibiotic resistance patterns of Streptococcus pneumoniae serotype 5: prior to PCV-13 vaccine introduction in Eastern Gambia. BMC infectious diseases. 2016;16:33.
69. Mackenzie G, Usuf E, Jasseh M, Nsekpong D, Ikumapayi N, Badji H, et al. Population-based surveillance for pneumonia, sepsis and meningitis in all ages in The Gambia: Implications for pneumococcal vaccine introduction and surveillance in Africa. International Journal of Infectious Diseases. 2010;14:e27-e8.
70. Darboe MK, Fulford AJ, Secka O, Prentice AM. The dynamics of nasopharyngeal streptococcus pneumoniae carriage among rural Gambian mother-infant pairs. BMC infectious diseases. 2010;10:195.
71. Swarthout TD, Fronterre C, Lourenço J, Obolski U, Gori A, Bar-Zeev N, et al. High residual carriage of vaccine-serotype *Streptococcus pneumoniae* after introduction of pneumococcal conjugate vaccine in Malawi. Nature communications. 2020;11(1):2222.
72. Kamng'ona AW, Hinds J, Bar-Zeev N, Gould KA, Chaguza C, Msefula C, et al. High multiple carriage and emergence of Streptococcus pneumoniae vaccine serotype variants in Malawian children. BMC infectious diseases. 2015;15:234.
73. Bar-Zeev N, Swarthout TD, Everett DB, Alaerts M, Msefula J, Brown C, et al. Impact and effectiveness of 13-valent pneumococcal conjugate vaccine on population incidence of vaccine and non-vaccine serotype invasive pneumococcal disease in Blantyre, Malawi, 2006–18: prospective observational time-series and case-control studies. The Lancet Global Health. 2021;9(7):e989-e98.
74. Heinsbroek E, Tafatatha T, Phiri A, Swarthout TD, Alaerts M, Crampin AC, et al. Pneumococcal carriage in households in Karonga District, Malawi, before and after introduction of 13-valent pneumococcal conjugate vaccination. Vaccine. 2018;36(48):7369-76.
75. Adebanjo T, Lessa FC, Mucavele H, Moiane B, Chauque A, Pimenta F, et al. Pneumococcal carriage and serotype distribution among children with and without pneumonia in Mozambique, 2014-2016. PloS one. 2018;13(6):e0199363.
76. Valenciano SJ, Moiane B, Lessa FC, Chaúque A, Massora S, Pimenta FC, et al. Effect of 10-Valent Pneumococcal Conjugate Vaccine on Streptococcus pneumoniae Nasopharyngeal Carriage Among Children Less Than 5 Years Old: 3 Years Post-10-Valent Pneumococcal Conjugate Vaccine Introduction in Mozambique. Journal of the Pediatric Infectious Diseases Society. 2021;10(4):448-56.
77. Massora S, Lessa FC, Moiane B, Pimenta FC, Mucavele H, Chaúque A, et al. Invasive disease potential of Streptococcus pneumoniae serotypes before and after 10-valent pneumococcal conjugate vaccine introduction in a rural area, southern Mozambique. Vaccine. 2019;37(51):7470-7.

78. Sigaúque B, Moiane B, Massora S, Pimenta F, Verani JR, Mucavele H, et al. Early Declines in Vaccine Type Pneumococcal Carriage in Children Less Than 5 Years Old After Introduction of 10-valent Pneumococcal Conjugate Vaccine in Mozambique. The Pediatric infectious disease journal. 2018;37(10):1054-60.
79. Sigaúque B, Verani JR, Massora S, Vubil D, Quintó L, Acácio S, et al. Burden of invasive pneumococcal disease among children in rural Mozambique: 2001-2012. PloS one. 2018;13(1):e0190687.
80. Vallès X, Flannery B, Roca A, Mandomando I, Sigaúque B, Sanz S, et al. Serotype distribution and antibiotic susceptibility of invasive and nasopharyngeal isolates of Streptococcus pneumoniae among children in rural Mozambique. Trop Med Int Health. 2006;11(3):358-66.
81. Løchen A, Truscott JE, Croucher NJ. Analysing pneumococcal invasiveness using Bayesian models of pathogen progression rates. PLOS Computational Biology. 2022;18(2):e1009389.
